# Neck pain care pathways and costs: association with the type of initial contact health care provider. A retrospective cohort study

**DOI:** 10.1101/2022.07.18.22277777

**Authors:** David Elton, Meng Zhang

**Affiliations:** UnitedHealth Group, Eden Prairie, Minnesota USA; Optum Labs, Eden Prairie, Minnesota USA

**Keywords:** Neck pain, pathway, guideline, initial contact, first provider, utilization, cost, value

## Abstract

**Background:** For individuals with neck pain (NP) the type of health care provider (HCP) initially contacted and subsequent services used are less well understood than for low back pain (LBP). The purpose of this retrospective observational study of administrative data was to examine the association between type of HCP initially contacted by individuals with NP, service utilization, and total episode cost.

**Methods:** A US national sample of NP episodes completed in 2017-2019 was analyzed. Separate analyses of a combined surgical and non-surgical (pooled) sample and a non-surgical sample were performed. Seventeen types of HCP initially contacted by an individual with NP was the primary independent variable. Dependent measures included rate and timing of use of fourteen types of health care services and total episode cost. A mixed effects model applied to pooled and non-surgical samples was used to test the association between initial contact HCP, total episode cost and rate of prescription opioid and NSAID use for NP.

**Results:** The study included 323,348 continuously insured individuals aged 18 years and older with 390,992 complete episodes of NP involving 321,538 HCPs and incurring $472,399,064 in expenditures. 53.0% of episodes had initial contact with a primary care or specialist HCP, with these episodes associated with higher rates of imaging, pharmaceutical, and interventional services. 40.4% of episodes had initial contact with a non-prescribing HCP, with these episodes associated with higher rates of non-pharmaceutical services. Chiropractors (DC) were the most common type of HCP initially contacted (38.5% of episodes) and were associated with the lowest adjusted total episode cost. Results were consistent for individuals experiencing single or multiple episodes during the study period.

**Conclusions:** This study of a large US cohort of commercially insured individuals with NP helps fill a knowledge gap regarding NP care pathway attributes. Like LBP, the treatment of NP is highly variable with the initial HCP selected by an individual with NP associated with differences in services received and episode cost. Initial contact with a non-prescribing HCP was associated with lower rates of imaging, pharmacology, and interventional services.

## Background

Neck pain (NP) has high prevalence and incidence.^1,2^ Annual United States (US) all-payer combined NP and low back pain (LBP) costs are estimated to be $134.5 billion, making spinal disorders the costliest condition type, more than all other musculoskeletal conditions combined ($129.8 billion) and more than the cost of diabetes ($111.2 billion).^3^ NP estimated years lived with disability (YLD) is also high ^1,4^, although YLD estimates may over or underrepresent actual NP disability.^5-7^

Among spinal disorders, the management of LBP benefits from the availability of high-quality, relatively homogenous clinical practice guidelines (CPGs)^8-12^ that describe a stepped approach to management. In the absence of red flags of significant pathology services progress from first-line non-pharmaceutical and non-interventional services to selective use of second- and third-line pharmaceutical and interventional services in cases not responding to first-line approaches.^8-12^

Compared to LBP, NP has relatively fewer high-quality CPGs available to inform decision-making, with methodological limitations and heterogeneity identified.^13^ Three recent NP CPGs, with relatively high Appraisal of Guidelines for Research and Evaluation (AGREE) II scores ^13^, corroborate a 2008 literature synthesis ^14^ and recommend an approach similar to what is described in LBP CPGs. In the absence of serious pathology initial management of NP should emphasize non-pharmaceutical and non-interventional approaches.^15-17^

Healthcare services with minimal to no beneficial impact on outcomes, or associated with potential harm, are described as low-value.^18,19^ Low-value care for the management of LBP has been well described^20-24^ with almost half of low-value spending in US health care ^25^ attributed to mismanagement of LBP. The heterogeneity in NP CPGs results in the magnitude of low-value care associated with NP being less well understood than for LBP.

The initial contact health care provider (HCP) has been used to evaluate CPG concordance and variation in service utilization and cost outcomes for LBP.^26,27^ A similar approach has been used for NP, and, like LBP, these studies have found that early access to non-pharmaceutical and non-interventional HCPs like chiropractors (DC), physical therapists (PT), and licensed acupuncturists (LAc) is associated with lower rates of opioid prescribing and beneficial impact on other measures.^28-31^

The aims of this study were to provide a comprehensive summary of HCPs initially contacted by individuals with NP and to examine the association between the type of initial contact HCP, service utilization, and cost of care for the treatment of NP in a US national sample of commercially insured adults. The hypothesis was that neck pain service utilization and total episode cost would vary based on the type of initial contact HCP.

## Methods

### Study design, population, setting and data sources

This retrospective cohort study of individuals seen by one or more HCPs for a complete episode of NP used methods similar to a recent LBP study.^32^ A comprehensive analytic database was created by linking multiple databases. An enrollee database included de-identified enrollment records, administrative claims data for all inpatient and outpatient services, and pharmacy prescriptions for enrollees from a single national commercial health insurer. A HCP database consisted of de-identified in- and out-of-network HCP demographic information and professional licensure status. ZIP code level population race and ethnicity data were obtained from the US Census Bureau^33^, adjusted gross income (AGI) data from the Internal Revenue Service^34^, socioeconomic Area Deprivation Index (ADI) data was obtained from the University of Wisconsin Neighborhood Atlas® database.^35^

The analysis does not include an adjustment for typical known and measurable confounders such as individual age, sex and co-morbidities^36,37^ using common yet potentially inadequate approaches such as propensity score matching^38^ due to the inability to control for unknown and unmeasurable confounders, and confounders of measurable confounders. Examples of the types of data not available include; HCP options convenient to an individual’s home, workplace or daily travel routes including public transportation if used, individual preferences for type of HCP including gender or racial concordance and specific services, recommendations from family or friends and influence of local HCP marketing efforts, perceived NP severity, combining a NP visit with a previously scheduled visit for another unrelated condition or annual PCP visit, anticipated potential out of pocket costs, and appointment availability within an individual’s timing expectations for HCPs meeting these and other criteria.^39^ As an alternative to blurring the line between association and causation through a process that simultaneously introduces distortion and complexity into dependent measures, this descriptive analysis includes actual measures of individual demographic and episodic characteristics, and associations, reported for each type of HCP initially selected by individuals with NP.

With study data de-identified or a Limited Data Set in compliance with the Health Insurance Portability and Accountability Act and customer requirements, the UnitedHealth Group Office of Human Research Affairs Institutional Review Board determined that this study was exempt from ethics review. The study was conducted and reported based on the Strengthening the Reporting of Observational Studies in Epidemiology (STROBE) guidelines (Supplement – STROBE Checklist).^40^

### Unit of analysis and cohort selection

The unit of analysis was episode of care, which has been reported as a valid measurement for comparison of HCPs based on cost of care.^41,42^ Administrative claims data were translated into episodes of care using Symmetry^®^ Episode Treatment Groups^®^ (ETG^®^) and Episode Risk Groups^®^ (ERG^®^) version 9.5 methodologies and definitions. An episode sequence cohort categorization model^32^ (Figure 1) was developed as it was possible for an individual to have multiple episodes of NP during the study period.

**Figure 1.**
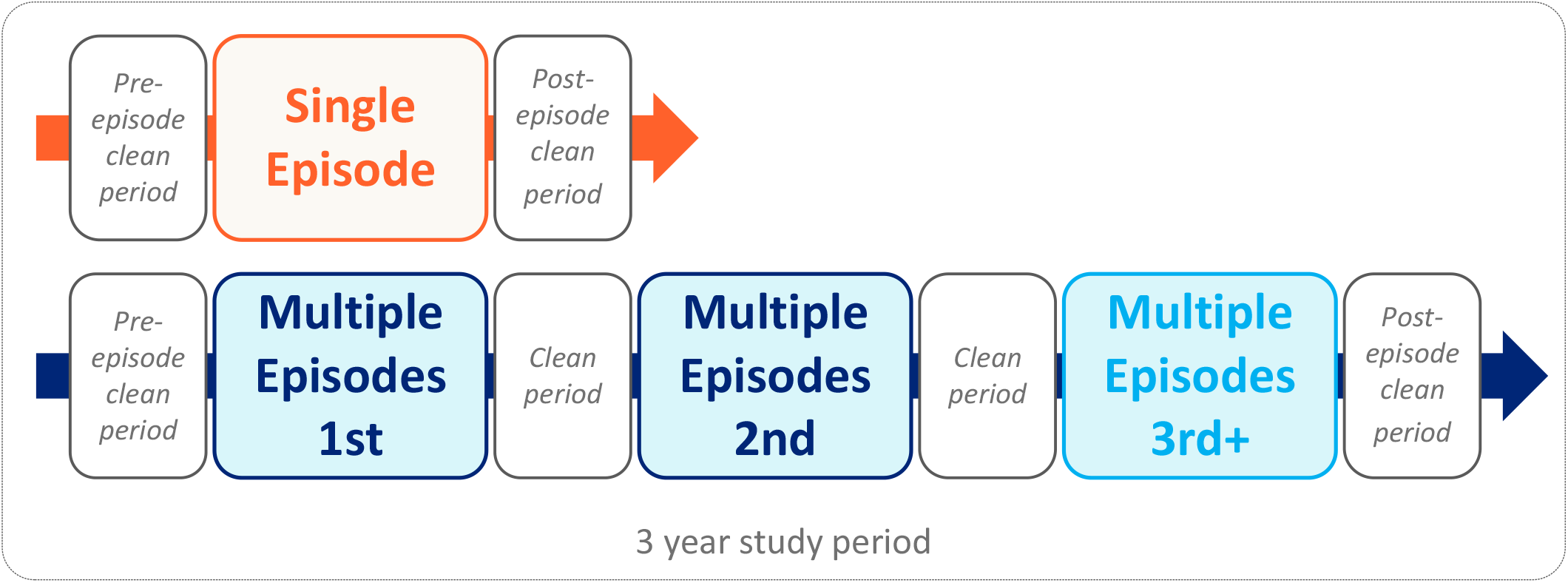
Episode sequence cohort and clean period conceptual model From “*Low back pain care pathways and costs: association with the type of initial contact health care provider. A longitudinal cohort study*” medRxiv 2022.06.17.22276443; doi: https://doi.org/10.1101/2022.06.17.22276443. Reprinted with permission.

The study evaluated complete episodes of neck pain defined by clean periods with no services provided for NP by any HCP for at least 91-days before the first NP visit and 61-days after the final NP visit. The number of days between the first and last visit was the episode duration. The three-year study period, coupled with the 150-day pre and post episode clean periods, was associated with 0.25% of episodes with a duration of greater than 2 years. Due to the unusual nature of NP involving continuous treatment for over 2 years, and incomplete sample of these episodes, these episodes were excluded from the analysis. NP episodes associated with diagnoses of malignant and non-malignant neoplasms, fractures and other spinal trauma, infection, congenital deformities and scoliosis, autoimmune disorders, osteoporosis, and advanced arthritis were also excluded.

The cohort consisted of individuals with continuous medical and pharmacy insurance coverage during the entire study period, who were 18 years of age and older, and who experienced a complete episode of NP beginning and ending during the calendar years 2017-2019. This timeframe was selected to align with the similar LBP study^32^, and as the three-year period before the influence of the COVID-19 epidemic on care patterns. Within the cohort, two samples were created: a non-surgical sample consisting of NP episodes without a surgical procedure and a pooled sample consisting of NP episodes with and without a surgical procedure. The pooled sample enabled a comprehensive examination of the associations between initial contact HCP and services provided for NP. With the potential for individuals of differing complexity to select different initial contact HCPs, analysis of the non-surgical sample sought to partially address this potential study limitation (Supplement – Cohort). The Results section describes the non-surgical sample with results for the pooled sample included in supplements.

### Variables

Python (Python Language Reference, Version 3.7.5., n.d.) was used for data preprocessing, table generation, and initial analyses. R (version 3.6.1) was used for linear mixed models regression. D’Agostino’s K-squared test was used for a goodness-of-fit analysis. Median, interquartile range (IQR), quartile 1 (Q1), and quartile 3 (Q3) were used to report non-normally distributed data.

The primary independent variable was the type of initial contact HCP, segmented into primary care, non-prescribing, specialist, and emergency/urgent care categories. 17 HCP types commonly contacted initially for an episode of NP were analyzed. All HCP types could be accessed directly without a referral. Excluded from the analysis were non-physician HCP types infrequently initially contacted by an individual with NP and HCPs for whom a type could not be identified, often an out-of-network HCP. Doctors of Osteopathy (DO) were included in the HCP type for which the DO was boarded except for DOs with evidence of billing an Osteopathic Manipulative Treatment (OMT) Current Procedural Terminology^*®*^ (CPT^*®*^) code who were separately reported. Family Practice, Internal Medicine, General Medicine, and Obstetrician-Gynecologist physician types were grouped into a Primary Care Provider (PCP) category. A

Nurse category was created, with nurse practitioner being the most common. For medical physician types not included in the other categories an “MD-other” category was created. For episodes involving multiple HCPs during the initial episode visit, the initial type of HCP was assigned using the hierarchy of emergency medicine/urgent care, primary care, non-prescriber, and specialist, as this order approximated the most likely experience of an individual with NP.

The rate and timing of use of 14 types of health care was the primary dependent variable. While NP CPGs do not group health care services into first-, second-, or third-line, to facilitate a comparison of the management of NP and LBP services were organized in a manner consistent with a previous LBP study.^32^ For each type of initial contact HCP, the percent of episodes including each of the 14 types of health care services was calculated based on a service being provided by any HCP that an individual saw during the complete episode of NP. The timing of when a service was first performed within an episode was reported using the median and interquartile range due to the data not being normally distributed.

Odds (OR), risk (RR) ratios, and associated 95% confidence intervals, were calculated for utilization of each service type. RR were reported as this is the measure more widely understood in associational analyses and due to the tendency for ORs to exaggerate risk in situations where an outcome is relatively common^43^. While DCs were the most common type of HCP initially contacted by individuals with NP, to remain consistent with a previous LBP study^32^ the RR baseline was episodes with a PCP as the initial contact HCP. In contrast to a previous LBP study^32^, and due to relative lack of uniform, high quality NP CPGs, the NP study does not include bivariate analyses compared with a PCP reference.

The secondary dependent variable was the total cost of care for all reimbursed services provided by any HCP. Total cost of care included services (e.g., evaluation, durable medical equipment, lab studies, etc.) not separately reported in the 14 categories of services described in the analysis. Cost information was not available for services for which an insurance claim was not submitted. Indirect costs associated with lost productivity or missed days at work were not included. Total episode cost was reported using the median and interquartile range due to the data not being normally distributed.

To test the association between type of initial HCP, total episode cost, opioid use and NSAID use a mixed effects model was developed and applied to both pooled and non-surgical samples. To remain consistent with a previous LBP study^32^, the initial type of HCP, individual age, sex, and ERG^®^ risk score were considered fixed effects, the intercept was PCPs initially contacted by male individuals of average cohort age with an ERG^®^ score of zero. To account for individual HCP influenced decision making and cost differences a unique identifier for the initial HCP was included as a random effect.

## Results

The non-surgical sample included 311,394 individuals associated with 374,403 complete NP episodes involving 286,761 unique HCPs. There were $221,821,643 in reimbursed health care expenditures with a median total cost per episode of $205 (IQR Q1, Q3 $81, $591) (Table 1a). Individuals were from all 50 states and some U.S. territories (Supplement 1).

**Table 1a.** Neck pain cohort characteristics.

|  | Pooled | Non-Surgical |
| --- | --- | --- |
| Individuals | 323348 | 311394 |
| Complete Episodes | 390992 | 374403 |
| Health Care Providers (HCP) | 321538 | 286761 |
| Total Episode Cost | 472399659 | 221821643 |
| Episodes - Median (Q1, Q3) |  |  |
| Total Episode Cost | \$224 (87, 691) | \$205 (81, 591) |
| Individuals - Median (Q1, Q3) |  |  |
| Age (years) | 45 (35, 54) | 45 (34, 54) |
| ERG<sup>®</sup> Risk Score | 1.5 (0.7, 3.0) | 1.5 (0.7, 2.9) |
| Episodes Per Individual | 1 (1, 1) | 1 (1, 1) |
| Individuals - % |  |  |
| Female | 59.9% | 60.1% |
| Non-Hispanic White | 48.7% | 48.4% |
| Hispanic | 5.7% | 5.7% |
| Non-Hispanic Black | 4.0% | 4.0% |
| Non-Hispanic Asian | 1.3% | 1.3% |
| Non-Hispanic Native American | 0.1% | 0.1% |
| Non-Hispanic Pacific Islander | 0.1% | 0.1% |
| Multiple | 5.5% | 5.5% |
| Other | 2.4% | 2.5% |
| Unknown | 32.1% | 32.3% |

82.8% of individuals, generating 68.5% of episodes, had a single complete NP episode during the study period. The actual pre- and post-episode clean periods were substantially longer than the ETG^®^ clean period definitions. For individuals with a single complete episode, the median pre-episode clean period was 650 days (Q1 438, Q3 865). For individuals with multiple episodes, the median pre-episode clean period before the first of multiple episodes was 373 days (Q1 212, Q3 558). The median number of days between sequential episodes was 189 days (Q1 116, Q3 316). The median post-episode clean period was 401 days (Q1 236, Q3 617) (Supplement 2).

DCs (38.5% of episodes) and PCPs (27.6%) were the most common types of HCP initially contacted by individuals with NP. Orthopedic surgeons (OS) (6.2%) were the most common type of specialist HCP initially contacted. The characteristics of individuals, episodes, and local population factors was variable for the different types of HCP initially contacted. DC, LAc, EM, and UC HCPs were initially contacted by younger individuals (median age 39-42) and with a lower ERG® risk score (median less than 1.5). LAc and PT HCPs were initially contacted by individuals from zip codes with lower levels of deprivation (ADI score less than 35) (Table 1b).

**Table 1b.**
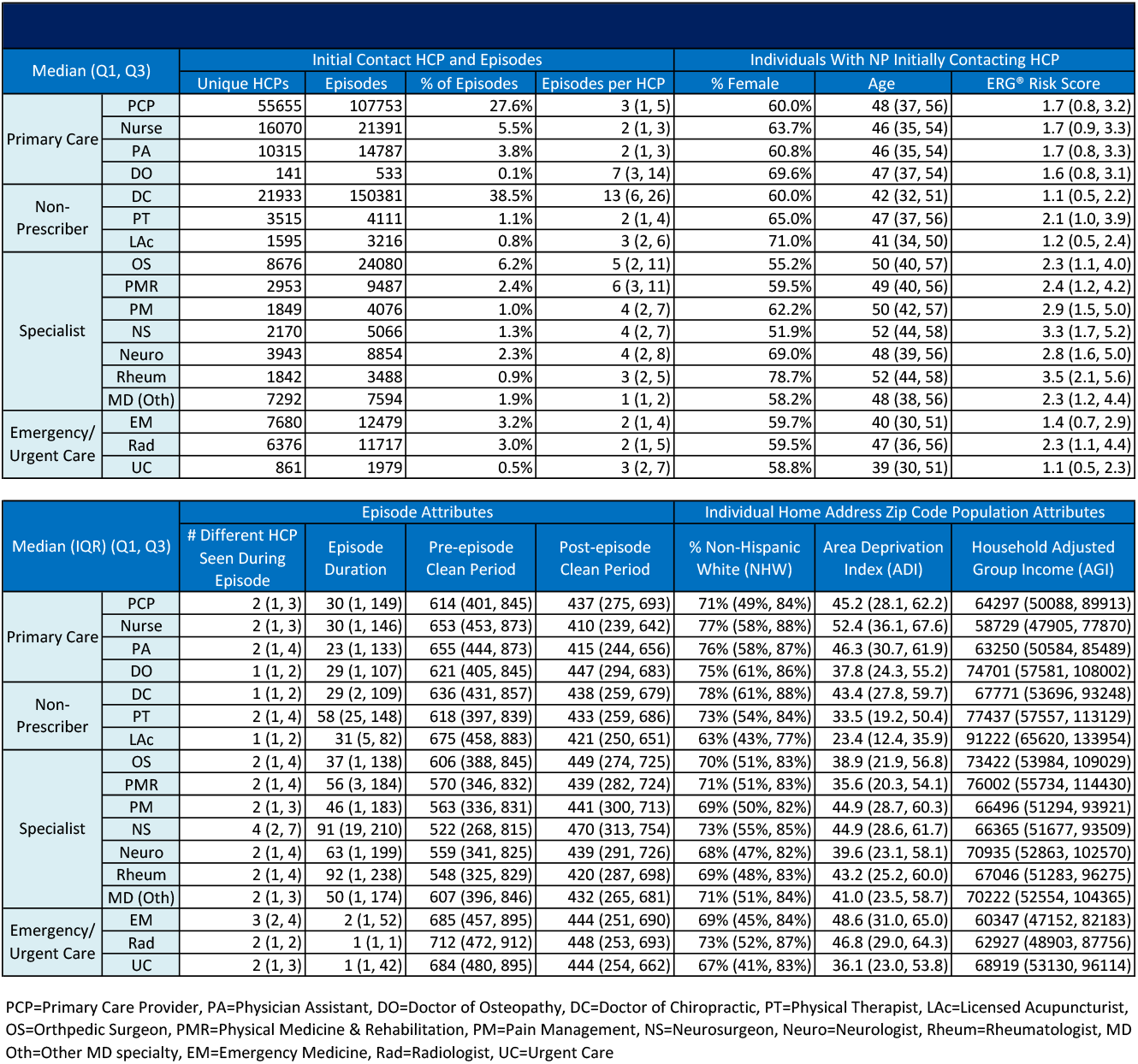
Episode, individual and population characteristics associated with type of health care provider (HCP) initially contacted by individuals with neck pain (NP)

The most frequently provided first-line services were chiropractic manipulation (40.2% of episodes), active care (21.5%), and passive therapy (18.9%). The most frequently provided second-line services were radiographs (26.5%), skeletal muscle relaxants (19.2%), and prescription NSAIDs (18.3%). The most frequently provided third-line services were prescription opioids (11.6%) and spinal injections (5.0%). 4.2% of episodes included spinal surgery. If a first- or second-line service was provided, other than MRI, the median timing was within the first 7 days of an episode, and most commonly on the initial visit (Table 1c).

**Table 1c.** Services provided for neck pain.

|  |  | Episodes Including Service | % of Episodes | Days Into Episode When Initially Provided<br>-<br>Median (IQR)(Q1, Q3) |
| --- | --- | --- | --- | --- |
| First Line | Manipulation - Chiropractic | 157371 | 40.2% | 0 (0) (0, 0) |
|  | Active Care | 83881 | 21.5% | 4 (30) (0, 30) |
|  | Passive Therapy | 73902 | 18.9% | 0 (20) (0, 20) |
|  | Manual Therapy | 61975 | 15.9% | 7 (37) (0, 37) |
|  | Acupuncture | 5080 | 1.3% | 0 (18) (0, 18) |
|  | Manipulation - Osteopathic | 4530 | 1.2% | 0 (11) (0, 11) |
| Second Line | Imaging - Radiography | 103447 | 26.5% | 0 (14) (0, 14) |
|  | Rx - Skeletal Muscle Relaxant | 75034 | 19.2% | 0 (16) (0, 16) |
|  | Rx - NSAID | 71442 | 18.3% | 0 (41) (0, 41) |
|  | Imaging - MRI | 43058 | 11.0% | 17 (60) (2, 62) |
| Third Line | Rx - Opioid | 45196 | 11.6% | 7 (69) (0, 69) |
|  | Spinal Injection | 19635 | 5.0% | 35 (93) (5, 98) |
|  | Spinal Surgery | 16589 | 4.2% | 46 (89) (15, 104) |
|  | Imaging - CT | 15753 | 4.0% | 0 (35) (0, 35) |

For HCP types initially contacted by individuals with NP, the percent of episodes including services categorized as first-, second- and third-line, and the timing of when these services were first introduced during an episode, was variable. Initial contact with primary care, specialist and emergency/urgent care HCPs was associated with pharmaceutical, imaging, and interventional services provided most often, and if provided, typically within the first 7 days of an episode and commonly during the initial visit. Initial contact with non-prescribing HCPs was associated with one or more first-line therapies provided during the initial visit. For individuals with NP initially contacting a non-prescribing HCP, pharmaceutical, imaging, and interventional services, other than radiography, are infrequently provided, and if provided, are introduced later in an episode. Tables 2a and 2b present these data for the non-surgical sample and single episode cohort.

**Table 2a.** Neck pain % of episodes including service by type of initial contact health care provider (HCP)

|  |  |  |  |  |  |  |  |  |  |  |  |  |  |  |  |  |  |  |  |  |
| --- | --- | --- | --- | --- | --- | --- | --- | --- | --- | --- | --- | --- | --- | --- | --- | --- | --- | --- | --- | --- |
| Non-Surgical Single Episode Cohort |  | Episodes |  | First Line |  |  |  |  |  |  | Second Line |  |  |  |  | Third Line |  |  |  |  |
|  |  | Count | % | Any | Manipulation - Chiropractic | Active Care | Passive Therapy | Manual Therapy | Acupuncture | Manipulation - Osteopathic | Any | Imaging - Radiography | Rx - NSAID | Rx - Skeletal Muscle Relaxant | Imaging - MRI | Any | Rx-Opioid | Spinal Injection | Spinal Surgery | Imaging-CT |
| Total |  | 256203 | 100.0% | 48.6% | 36.8% | 20.8% | 18.1% | 15.2% | 1.3% | 1.1% | 53.6% | 28.9% | 18.2% | 20.5% | 9.2% | 15.2% | 9.8% | 2.4% | 0.0% | 4.4% |
| Primary Care | PCP | 74620 | 26.8% | 22.2% | 8.3% | 13.9% | 8.1% | 11.2% | 0.4% | 2.7% | 67.0% | 27.4% | 29.8% | 35.4% | 9.0% | 19.0% | 14.9% | 2.4% | 0.0% | 3.2% |
|  | Nurse | 15059 | 5.4% | 21.7% | 10.8% | 13.7% | 9.1% | 9.8% | 0.3% | 0.6% | 72.7% | 28.8% | 30.9% | 42.3% | 8.5% | 20.4% | 13.6% | 4.2% | 0.0% | 4.6% |
|  | PA | 10563 | 3.8% | 20.7% | 8.7% | 14.5% | 7.2% | 11.5% | 0.2% | 0.5% | 76.1% | 32.8% | 29.8% | 42.5% | 10.6% | 21.8% | 14.4% | 3.6% | 0.0% | 5.8% |
|  | DO | 344 | 0.1% | 68.6% | 2.6% | 17.4% | 8.4% | 10.2% | 0.0% | 62.2% | 22.4% | 10.2% | 6.4% | 7.0% | 5.5% | 11.6% | 6.1% | 5.2% | 0.0% | 1.5% |
|  | All | 100586 | 36.2% | 22.1% | 8.7% | 13.9% | 8.1% | 11.1% | 0.4% | 2.3% | 68.7% | 28.1% | 29.9% | 37.1% | 9.1% | 19.5% | 14.6% | 2.8% | 0.0% | 3.7% |
| Non-Prescriber | DC | 91026 | 32.8% | 94.6% | 90.1% | 30.0% | 35.2% | 18.9% | 0.8% | 0.1% | 26.2% | 20.5% | 4.6% | 3.8% | 2.1% | 4.4% | 3.2% | 0.8% | 0.0% | 0.6% |
|  | PT | 2805 | 1.0% | 97.4% | 10.6% | 93.8% | 31.7% | 80.1% | 2.0% | 0.7% | 40.1% | 21.4% | 10.1% | 9.2% | 16.9% | 10.9% | 5.8% | 3.4% | 0.0% | 3.0% |
|  | LAc | 2229 | 0.8% | 97.7% | 11.4% | 17.0% | 40.2% | 43.6% | 89.6% | 0.8% | 10.2% | 5.3% | 2.9% | 2.5% | 3.1% | 4.3% | 2.9% | 1.4% | 0.0% | 0.3% |
|  | All | 96060 | 34.6% | 94.7% | 86.0% | 31.6% | 35.2% | 21.3% | 2.9% | 0.2% | 26.3% | 20.2% | 4.7% | 3.9% | 2.6% | 4.6% | 3.3% | 0.9% | 0.0% | 0.7% |
| Specialist | OS | 15720 | 5.7% | 27.5% | 5.2% | 23.8% | 10.8% | 19.5% | 0.4% | 0.6% | 84.6% | 66.8% | 23.0% | 13.1% | 24.1% | 15.1% | 9.6% | 4.5% | 0.0% | 2.7% |
|  | PMR | 5749 | 2.1% | 33.3% | 6.3% | 28.4% | 12.3% | 22.8% | 1.6% | 2.0% | 65.5% | 37.7% | 20.1% | 14.9% | 23.6% | 25.8% | 15.7% | 11.4% | 0.0% | 2.3% |
|  | PM | 2037 | 0.7% | 11.5% | 1.6% | 9.4% | 3.9% | 7.7% | 0.2% | 0.8% | 47.8% | 13.1% | 13.8% | 18.4% | 16.8% | 41.4% | 30.8% | 13.9% | 0.0% | 1.7% |
|  | NS | 2467 | 0.9% | 21.3% | 3.8% | 18.2% | 8.5% | 14.9% | 0.2% | 0.2% | 75.7% | 40.5% | 17.3% | 15.5% | 43.8% | 22.7% | 14.1% | 4.7% | 0.0% | 8.5% |
|  | Neuro | 5061 | 1.8% | 19.6% | 5.5% | 15.5% | 7.7% | 12.6% | 0.5% | 0.4% | 65.5% | 13.4% | 18.8% | 26.3% | 32.2% | 19.0% | 10.3% | 7.2% | 0.0% | 3.9% |
|  | Rheu | 2019 | 0.7% | 18.0% | 8.0% | 12.0% | 7.0% | 10.2% | 0.6% | 0.5% | 74.7% | 39.0% | 26.4% | 26.4% | 14.1% | 21.4% | 15.3% | 5.4% | 0.0% | 2.8% |
|  | MD (Oth) | 4772 | 1.7% | 24.7% | 13.7% | 12.8% | 8.8% | 10.5% | 0.9% | 2.6% | 45.4% | 18.8% | 20.5% | 17.2% | 9.5% | 28.4% | 22.6% | 2.3% | 0.0% | 5.4% |
|  | All | 37825 | 13.6% | 25.2% | 6.4% | 20.2% | 9.6% | 16.5% | 0.6% | 1.0% | 71.1% | 43.1% | 21.0% | 16.8% | 23.6% | 21.2% | 14.0% | 6.2% | 0.0% | 3.5% |
| Emergency / Urgent Care | EM | 10565 | 3.8% | 11.1% | 4.6% | 7.7% | 4.8% | 6.2% | 0.2% | 0.3% | 70.5% | 35.0% | 32.4% | 42.1% | 5.7% | 41.2% | 15.5% | 1.3% | 0.0% | 29.6% |
|  | Rad | 9526 | 3.4% | 3.9% | 0.6% | 3.4% | 1.3% | 2.7% | 0.1% | 0.0% | 80.2% | 60.2% | 1.5% | 1.5% | 24.9% | 25.6% | 0.8% | 0.7% | 0.0% | 24.5% |
|  | UC | 1641 | 0.6% | 13.0% | 6.0% | 7.7% | 5.1% | 7.1% | 0.2% | 1.0% | 66.2% | 30.9% | 25.5% | 40.0% | 5.1% | 12.6% | 9.3% | 1.2% | 0.0% | 3.0% |
|  | All | 21732 | 7.8% | 8.1% | 3.0% | 5.8% | 3.3% | 4.7% | 0.1% | 0.2% | 74.5% | 45.8% | 18.3% | 24.1% | 14.1% | 32.2% | 8.6% | 1.0% | 0.0% | 25.3% |

PCP=Primary Care Provider, PA=Physician Assistant, DO=Doctor of Osteopathy, DC=Doctor of Chiropractic, PT=Physical Therapist, LAc=Licensed Acupuncturist, OS=Orthpedic Surgeon, PMR=Physical Medicine & Rehabilitation, PM=Pain Management, NS=Neurosurgeon, Neuro=Neurologist, Rheum=Rheumatologist, MD Oth=Other MD specialty, EM=Emergency Medicine, Rad=Radiologist, UC=Urgent Care

**Table 2b.** Neck pain # of days into episode when service initially provided by type of initial contact health care provider (HCP)

|  |  |  |  |  |  |  |  |  |
| --- | --- | --- | --- | --- | --- | --- | --- | --- |
| Non-Surgical Single Episode Cohort<br>Median (Q1, Q3) |  | First Line |  |  |  |  |  |  |
|  |  | Any | Manipulation - Chiropractic | Active Care | Passive Therapy | Manual Therapy | Acupuncture | Manipulation - Osteopathic |
| Primary Care | PCP | 18 (3, 58) | 39 (9, 99) | 21 (7, 58) | 30 (8, 81) | 21 (7, 58) | 29 (2, 88) | 0 (0, 0) |
|  | Nurse | 18 (4, 57) | 25 (5, 73) | 20 (5, 52) | 21 (5, 64) | 21 (6, 55) | 44 (16, 103) | 5 (0, 26) |
|  | PA | 20 (6, 58) | 29 (7, 84) | 19 (7, 53) | 27 (9, 72) | 20 (8, 54) | 47 (38, 106) | 16 (5, 57) |
|  | DO | 0 (0, 0) | 55 (9, 118) | 8 (0, 41) | 0 (0, 29) | 25 (8, 58) | N/A | 0 (0, 0) |
|  | All | 18 (3, 57) | 35 (8, 91) | 21 (7, 57) | 28 (7, 77) | 21 (7, 57) | 34 (4, 92) | 0 (0, 0) |
| Non-Prescriber | DC | 0 (0, 0) | 0 (0, 0) | 0 (0, 5) | 0 (0, 2) | 0 (0, 7) | 7 (0, 56) | 41 (6, 110) |
|  | PT | 0 (0, 0) | 38 (11, 60) | 0 (0, 2) | 4 (0, 22) | 0 (0, 6) | 28 (1, 100) | 38 (8, 108) |
|  | LAc | 0 (0, 0) | 36 (7, 59) | 4 (0, 36) | 0 (0, 0) | 0 (0, 2) | 0 (0, 0) | 28 (0, 56) |
|  | All | 0 (0, 0) | 0 (0, 0) | 0 (0, 5) | 0 (0, 2) | 0 (0, 7) | 0 (0, 0) | 38 (6, 98) |
| Specialist | OS | 14 (6, 36) | 41 (12, 104) | 14 (6, 35) | 20 (8, 49) | 15 (7, 35) | 61 (16, 120) | 0 (0, 34) |
|  | PMR | 11 (1, 31) | 24 (2, 93) | 12 (3, 31) | 18 (4, 46) | 14 (4, 34) | 0 (0, 37) | 0 (0, 21) |
|  | PM | 4 (0, 34) | 24 (0, 72) | 7 (0, 34) | 16 (0, 48) | 10 (0, 42) | 77 (38, 77) | 0 (0, 0) |
|  | NS | 28 (10, 58) | 61 (16, 140) | 28 (12, 55) | 32 (15, 69) | 29 (13, 58) | 99 (46, 260) | 82 (63, 116) |
|  | Neuro | 30 (10, 77) | 56 (22, 128) | 31 (11, 78) | 39 (15, 95) | 30 (11, 75) | 84 (31, 109) | 55 (4, 96) |
|  | Rheu | 45 (15, 109) | 63 (18, 125) | 49 (17, 122) | 72 (24, 138) | 48 (18, 112) | 18 (14, 43) | 18 (5, 89) |
|  | MD (Oth) | 38 (8, 93) | 49 (16, 102) | 51 (18, 118) | 54 (20, 112) | 50 (17, 119) | 59 (35, 79) | 0 (0, 0) |
|  | All | 17 (5, 50) | 46 (12, 107) | 17 (6, 48) | 24 (8, 65) | 19 (7, 49) | 37 (2, 95) | 0 (0, 23) |
| Emergency/<br>Urgent Care | EM | 20 (8, 45) | 17 (5, 53) | 24 (12, 48) | 24 (9, 51) | 26 (12, 47) | 22 (5, 37) | 13 (1, 66) |
|  | Rad | 19 (7, 37) | 29 (6, 65) | 20 (8, 37) | 24 (11, 43) | 21 (10, 41) | 33 (19, 71) | 60 (29, 93) |
|  | UC | 17 (6, 37) | 17 (6, 38) | 22 (10, 40) | 22 (7, 42) | 21 (10, 33) | 13 (7, 50) | 0 (0, 14) |
|  | All | 20 (7, 42) | 18 (5, 53) | 22 (11, 43) | 24 (9, 48) | 24 (11, 43) | 22 (7, 56) | 9 (0, 53) |

| Non-Surgical Single Episode Cohort<br>Median (Q1, Q3) |  | Second Line |  |  |  |  | Third Line |  |  |  |
| --- | --- | --- | --- | --- | --- | --- | --- | --- | --- | --- |
|  |  | Any | Imaging - Radiography | Rx - NSAID | Rx - Skeletal Muscle Relaxant | Imaging - MRI | Any | Rx-Opioid | Spinal Injection | Imaging-CT |
| Primary Care | PCP | 0 (0, 0) | 0 (0, 21) | 0 (0, 8) | 0 (0, 0) | 27 (8, 83) | 0 (0, 39) | 0 (0, 38) | 21 (0, 97) | 5 (0, 51) |
|  | Nurse | 0 (0, 0) | 0 (0, 18) | 0 (0, 5) | 0 (0, 0) | 28 (7, 83) | 1 (0, 34) | 2 (0, 48) | 13 (0, 56) | 0 (0, 25) |
|  | PA | 0 (0, 0) | 0 (0, 6) | 0 (0, 3) | 0 (0, 0) | 21 (6, 69) | 0 (0, 20) | 0 (0, 28) | 14 (0, 75) | 0 (0, 8) |
|  | DO | 4 (0, 30) | 4 (0, 26) | 30 (0, 89) | 4 (0, 26) | 21 (8, 28) | 13 (0, 48) | 29 (0, 54) | 10 (0, 41) | 0 (0, 26) |
|  | All | 0 (0, 0) | 0 (0, 19) | 0 (0, 7) | 0 (0, 0) | 27 (8, 80) | 0 (0, 35) | 0 (0, 38) | 16 (0, 82) | 1 (0, 36) |
| Non-Prescriber | DC | 0 (0, 22) | 0 (0, 0) | 77 (25, 155) | 50 (12, 133) | 50 (17, 121) | 53 (14, 133) | 66 (21, 153) | 35 (7, 97) | 29 (0, 100) |
|  | PT | 14 (0, 52) | 9 (0, 51) | 42 (12, 108) | 28 (7, 82) | 35 (4, 71) | 26 (3, 82) | 30 (8, 94) | 51 (16, 116) | 2 (0, 58) |
|  | LAc | 36 (8, 104) | 42 (13, 97) | 47 (16, 139) | 33 (8, 96) | 42 (17, 172) | 68 (23, 130) | 88 (24, 170) | 68 (30, 94) | 27 (12, 138) |
|  | All | 0 (0, 26) | 0 (0, 0) | 75 (23, 153) | 48 (12, 131) | 46 (14, 112) | 51 (13, 130) | 64 (20, 151) | 38 (7, 97) | 26 (0, 97) |
| Specialist | OS | 0 (0, 0) | 0 (0, 0) | 0 (0, 26) | 0 (0, 26) | 9 (2, 28) | 7 (0, 56) | 7 (0, 64) | 21 (0, 64) | 13 (0, 64) |
|  | PMR | 0 (0, 5) | 0 (0, 0) | 5 (0, 63) | 0 (0, 38) | 10 (1, 35) | 2 (0, 36) | 2 (0, 38) | 14 (0, 65) | 20 (0, 93) |
|  | PM | 0 (0, 21) | 0 (0, 27) | 7 (0, 70) | 0 (0, 28) | 12 (0, 42) | 0 (0, 13) | 0 (0, 11) | 6 (0, 42) | 41 (1, 124) |
|  | NS | 0 (0, 6) | 0 (0, 16) | 30 (2, 106) | 12 (0, 80) | 1 (0, 16) | 16 (0, 64) | 20 (1, 82) | 64 (20, 98) | 13 (0, 55) |
|  | Neuro | 0 (0, 15) | 19 (0, 78) | 14 (0, 82) | 0 (0, 18) | 9 (0, 28) | 7 (0, 68) | 22 (0, 96) | 8 (0, 74) | 7 (0, 40) |
|  | Rheu | 0 (0, 2) | 0 (0, 29) | 0 (0, 34) | 0 (0, 28) | 42 (7, 130) | 9 (0, 77) | 10 (0, 82) | 9 (0, 90) | 46 (10, 100) |
|  | MD (Oth) | 1 (0, 59) | 23 (0, 92) | 2 (0, 76) | 9 (0, 78) | 44 (7, 125) | 0 (0, 20) | 0 (0, 19) | 52 (5, 126) | 0 (0, 36) |
|  | All | 0 (0, 1) | 0 (0, 0) | 1 (0, 49) | 0 (0, 36) | 9 (0, 33) | 2 (0, 45) | 2 (0, 49) | 17 (0, 70) | 11 (0, 64) |
| Emergency/<br>Urgent Care | EM | 0 (0, 0) | 0 (0, 0) | 0 (0, 2) | 0 (0, 1) | 13 (0, 48) | 0 (0, 0) | 0 (0, 4) | 2 (0, 58) | 0 (0, 0) |
|  | Rad | 0 (0, 0) | 0 (0, 0) | 9 (0, 60) | 1 (0, 22) | 0 (0, 0) | 0 (0, 0) | 18 (1, 64) | 35 (6, 70) | 0 (0, 0) |
|  | UC | 0 (0, 0) | 0 (0, 0) | 0 (0, 0) | 0 (0, 0) | 22 (8, 74) | 0 (0, 14) | 0 (0, 18) | 14 (0, 71) | 0 (0, 0) |
|  | All | 0 (0, 0) | 0 (0, 0) | 0 (0, 2) | 0 (0, 1) | 0 (0, 9) | 0 (0, 0) | 0 (0, 6) | 14 (0, 63) | 0 (0, 0) |

PCP=Primary Care Provider, PA=Physician Assistant, DO=Doctor of Osteopathy, DC=Doctor of Chiropractic, PT=Physical Therapist, LAc=Licensed Acupuncturist, OS=Orthpedic Surgeon, PMR=Physical Medicine & Rehabilitation, PM=Pain Management, NS=Neurosurgeon, Neuro=Neurologist, Rheum=Rheumatologist, MD Oth=Other MD specialty, EM=Emergency Medicine, Rad=Radiologist, UC=Urgent Care

Figure 2 compares the different patterns in service use for PCPs, DCs, and OSs initially contacted by an individual with NP. Supplement – Care Pathways illustrates these patterns for additional types of HCP.

**Figure 2.**
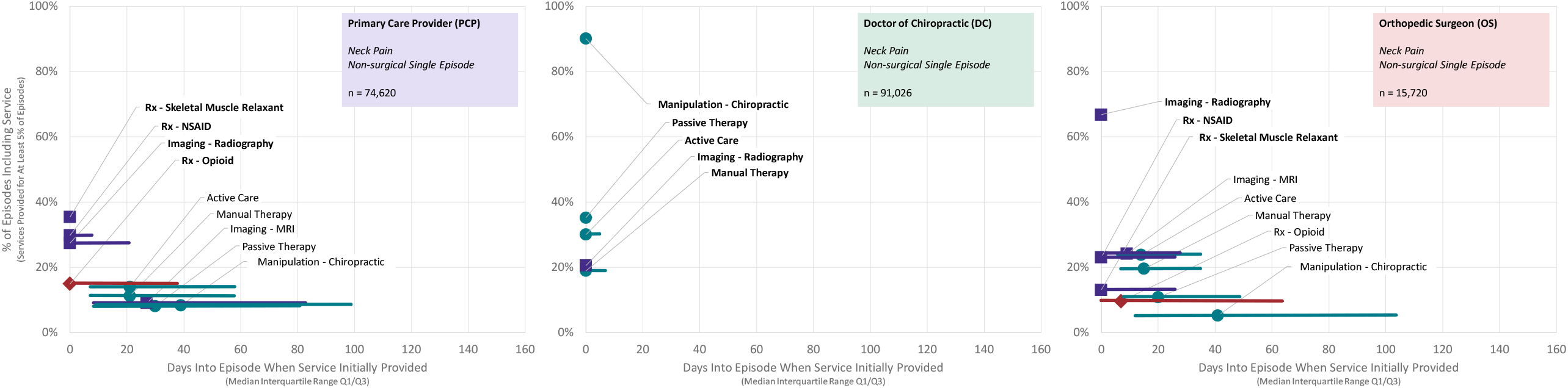
Rate and timing of use of health care services for individuals with neck pain initially contacting a primary care provider, chiropractor, and orthopedic surgeon - non-surgical single episode cohort

Figure 3 presents the RR comparing each type of initial contact HCP with the PCP reference.

**Figure 3.**
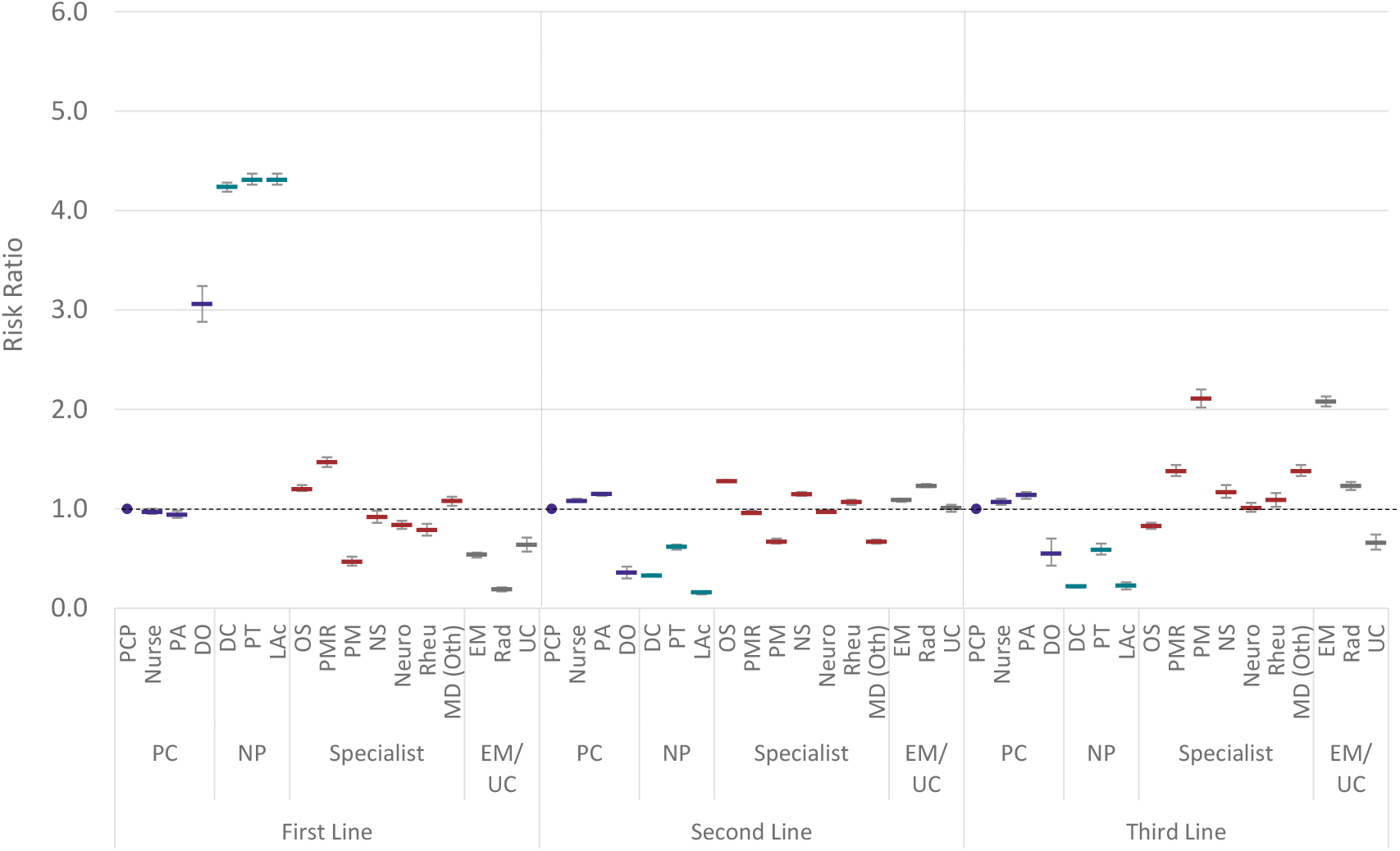
Risk ratio and 95% confidence interval for non-surgical neck pain episodic service exposure by initial contact health care provider compared to PCP reference. PC=Primary Care, NP=Non-Prescriber, EM/UC=Emergency Medicine/Urgent Care, PCP=Primary Care Provider; PA=Physician Assistant; DO=Doctor of Osteopathy; DC=Doctor of Chiropractic; PT=Physical Therapist; LAc=Licensed Acupuncturist; OS=Orthopedic Surgeon; PMR=Physical Medicine and Rehabilitation; PM=Pain Management, NS=Neurosurgeon; Neuro=Neurologist; Rheum=Rheumatologist; MD (Oth)=Other medical physician specialties, EM=Emergency Medicine; Rad=Radiologist; UC=Urgent Care

Supplement 3 presents the percent of episodes including each type of service for the overall non-surgical sample and Supplement 4 presents the results for the overall pooled sample. RRs are presented for the non-surgical (Supplement – 3a) and pooled (Supplement – 4a) samples. Timing data is presented for the overall non-surgical (Supplement 5) and pooled (Supplement 6) samples. Due to the volume of data associated with replicating Tables 2a and 2b, this information is not separately reported for all episode sequence cohorts.

There was a high degree of homogeneity in the distribution of initial contact HCP among episode sequence cohorts. For each type of initial contact HCP there was also a high degree of homogeneity among episode sequence cohorts in the rate of service use (Figure 4)

**Figure 4.**
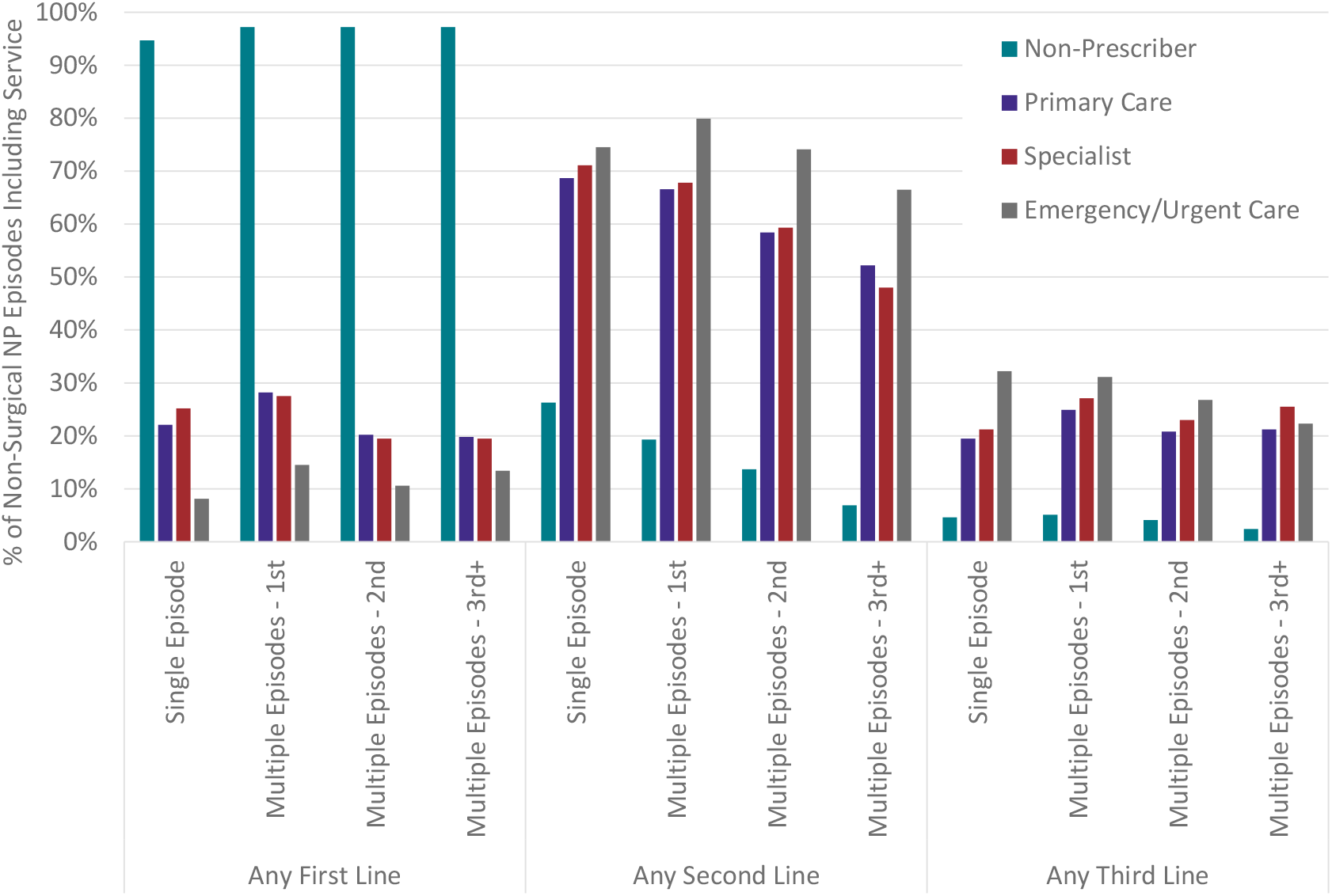
Percent of neck pain episodes including any first-, second- or third-line service by episode sequence cohort and type of initial contact health care provider

Total episode cost was variable for each type of HCP initially contacted by an individual with NP (Table 2c). Supplement 7 presents this same data for the pooled sample.

**Table 2c.** Neck pain total episode cost by episode sequence cohort.

| Non-surgical episodes<br>Median (Q1, Q3) |  | Total | Single episode | Multiple episodes |  |  |
| --- | --- | --- | --- | --- | --- | --- |
|  |  |  |  | First | Second | Third+ |
| Primary Care | PCP | \$153 (54, 466) | \$157 (60, 471) | \$185 (62, 622) | \$117 (25, 351) | \$94 (21, 250) |
| | Nurse | \$151 (36, 539) | \$159 (41, 557) | \$192 (47, 756) | \$96 (16, 353) | \$92 (20, 252) |
| | PA | \$197 (49, 680) | \$205 (58, 686) | \$240 (63, 917) | \$124 (18, 493) | \$94 (17, 292) |
| | DO | \$228 (105, 537) | \$248 (107, 586) | \$219 (82, 498) | \$196 (115, 453) | \$71 (69, 113) |
| | All | \$157 (51, 495) | \$162 (57, 503) | \$191 (60, 664) | \$115 (22, 362) | \$93 (20, 254) |
| Non-Prescriber | DC | \$162 (65, 400) | \$180 (70, 450) | \$165 (66, 385) | \$130 (58, 305) | \$90 (50, 180) |
| | PT | \$640 (323, 1334) | \$612 (306, 1317) | \$712 (382, 1384) | \$683 (350, 1421) | \$644 (254, 1026) |
| | LAc | \$330 (153, 707) | \$330 (153, 707) | \$388 (157, 888) | \$320 (148, 630) | \$276 (160, 552) |
| | All | \$167 (65, 420) | \$189 (77, 480) | \$170 (69, 405) | \$135 (60, 325) | \$90 (50, 187) |
| Specialist | OS | \$301 (130, 898) | \$296 (131, 895) | \$424 (161, 1163) | \$249 (107, 738) | \$216 (91, 647) |
| | PMR | \$496 (176, 1318) | \$508 (186, 1322) | \$604 (196, 1577) | \$392 (134, 1058) | \$276 (76, 991) |
| | PM | \$266 (100, 833) | \$302 (110, 926) | \$284 (105, 807) | \$194 (82, 620) | \$165 (80, 408) |
| | NS | \$624 (222, 1669) | \$672 (237, 1707) | \$775 (256, 1945) | \$438 (167, 1196) | \$254 (131, 858) |
| | Neuro | \$393 (114, 1117) | \$454 (138, 1187) | \$428 (125, 1221) | \$257 (78, 780) | \$124 (42, 311) |
| | Rheu | \$243 (67, 791) | \$227 (59, 786) | \$306 (102, 926) | \$242 (72, 761) | \$154 (38, 470) |
| | MD (Oth) | \$139 (25, 520) | \$149 (27, 590) | \$166 (33, 655) | \$100 (16, 326) | \$91 (22, 279) |
| | All | \$321 (113, 989) | \$334 (119, 1012) | \$403 (136, 1200) | \$242 (84, 764) | \$167 (62, 486) |
| Emergency/<br>Urgent Care | EM | \$861 (235, 1952) | \$898 (248, 1975) | \$840 (239, 2062) | \$402 (52, 1569) | \$319 (18, 1083) |
| | Rad | \$182 (67, 623) | \$173 (66, 583) | \$241 (80, 910) | \$214 (69, 702) | \$192 (48, 797) |
| | UC | \$205 (110, 380) | \$203 (111, 370) | \$241 (92, 530) | \$215 (73, 449) | \$120 (18, 364) |
| | All | \$356 (106, 1315) | \$362 (110, 1335) | \$420 (108, 1364) | \$276 (65, 927) | \$221 (37, 925) |

PCP=Primary Care Provider, PA=Physician Assistant, DO=Doctor of Osteopathy, DC=Doctor of Chiropractic, PT=Physical Therapist, LAc=Licensed Acupuncturist, OS=Orthopedic Surgeon, PMR=Physical Medicine & Rehabilitation, PM=Pain Management, NS=Neurosurgeon, Neuro=Neurologist, Rheum=Rheumatologist, MD Oth=Other MD specialty, EM=Emergency Medicine, Rad=Radiologist, UC=Urgent Care

With adjustment for covariates using a mixed effects model virtually all HCPs were associated with significantly different total episode cost, opioid use and NSAID use than the PCP reference group (Table 3) (Supplement 8).

**Table 3.** Neck pain mixed effects model - Non-surgical Sample.

| Features | Total Episode Cost | % of Episodes With Opioid | % of Episodes With NSAID |
| --- | --- | --- | --- |
| PCP - (Intercept) | 447 (432, 462) | 0.137 (0.134, 0.139) | 0.301 (0.298, 0.304) |
| Nurse | 80 (52, 109) | -0.012 (-0.017, -0.007) | 0.014 (0.007, 0.020) |
| PA | 144 (110, 178) | -0.005 (-0.011, 0.001) | 0.002 (-0.005, 0.009) |
| DO | -47 (-259, 164) | -0.102 (-0.139, -0.065) | -0.238 (-0.284, -0.192) |
| DC | -132 (-152, -112) | -0.120 (-0.123, -0.116) | -0.258 (-0.262, -0.254) |
| PT | 571 (509, 633) | -0.107 (-0.118, -0.096) | -0.203 (-0.217, -0.189) |
| LAc | 151 (78, 225) | -0.122 (-0.135, -0.109) | -0.274 (-0.290, -0.258) |
| OS | 242 (209, 274) | -0.059 (-0.064, -0.053) | -0.055 (-0.062, -0.047) |
| PMR | 649 (597, 700) | 0.013 (0.004, 0.022) | -0.112 (-0.123, -0.101) |
| PM | 329 (256, 402) | 0.128 (0.115, 0.141) | -0.178 (-0.193, -0.162) |
| NS | 767 (700, 834) | -0.041 (-0.053, -0.029) | -0.151 (-0.166, -0.137) |
| Neuro | 473 (425, 521) | -0.067 (-0.076, -0.059) | -0.125 (-0.135, -0.114) |
| Rheum | 454 (383, 524) | -0.038 (-0.051, -0.026) | -0.040 (-0.056, -0.025) |
| MD (Oth) | 313 (270, 356) | 0.051 (0.043, 0.059) | -0.103 (-0.113, -0.094) |
| EM | 979 (942, 1015) | 0.005 (-0.001, 0.012) | 0.020 (0.012, 0.028) |
| Rad | 46 (7, 85) | -0.168 (-0.175, -0.161) | -0.303 (-0.312, -0.295) |
| UC | 77 (-20, 174) | -0.043 (-0.060, -0.026) | -0.013 (-0.034, 0.008) |
| Individual Age | 3 (3, 4) | 0.001 (0.001, 0.001) | 0.000 (0.000, 0.001) |
| Individual Gender | 6 (-5, 17) | -0.000 (-0.002, 0.001) | 0.003 (0.001, 0.005) |
| ERG® Risk Score | 45 (43, 47) | 0.011 (0.011, 0.011) | 0.003 (0.003, 0.003) |

PCP=Primary Care Provider, PA=Physician Assistant, DO=Doctor of Osteopathy, DC=Doctor of Chiropractic, PT=Physical Therapist, LAc=Licensed Acupuncturist, OS=Orthpedic Surgeon, PMR=Physical Medicine & Rehabilitation, PM=Pain Management, NS=Neurosurgeon, Neuro=Neurologist, Rheum=Rheumatologist, MD Oth=Other MD specialty, EM=Emergency Medicine, Rad=Radiologist, UC=Urgent Care
Values in red were not statistically significant (p > 0.05) compared to intercept
Values in black were statistically significant (p > 0.05) compared to intercept

DCs had significantly lower total episode cost with most specialist HCPs having significantly higher total episode cost compared to the PCP reference group (Figure 5).

**Figure 5.**
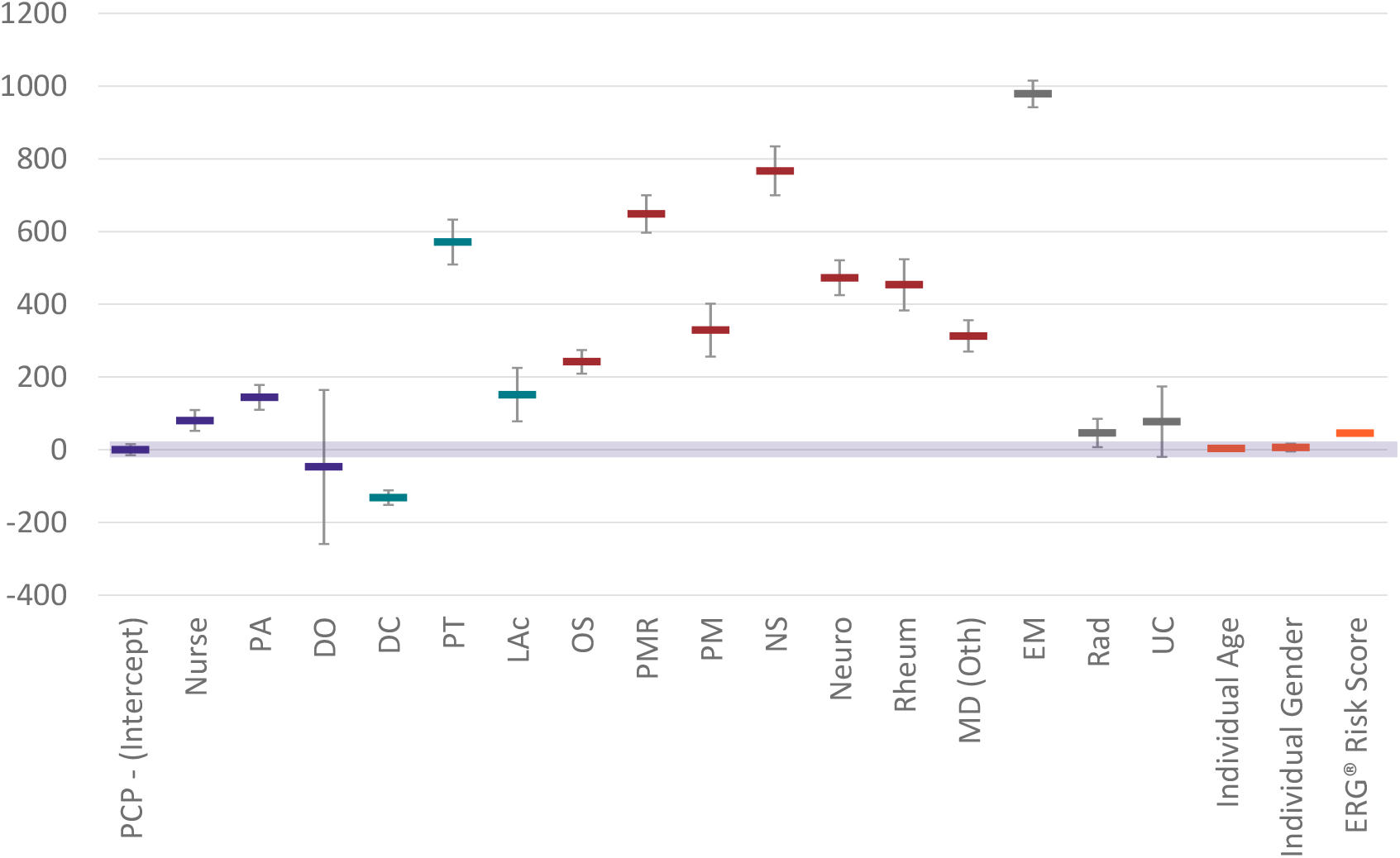
Neck pain non-surgical total episode cost regression estimates and 95% confidence intervals by type of initial contact HCP (0 Intercept = PCP reference group). PCP=Primary Care Provider; PA=Physician Assistant; DO=Doctor of Osteopathy; DC=Doctor of Chiropractic; PT=Physical Therapist; LAc=Licensed Acupuncturist; OS=Orthopedic Surgeon; PMR=Physical Medicine and Rehabilitation; PM=Pain Management, NS=Neurosurgeon; Neuro=Neurologist; Rheum=Rheumatologist; MD (Oth)=Other medical physician specialties, EM=Emergency Medicine; Rad=Radiologist; UC=Urgent Care

## Discussion

This study provides a comprehensive analysis of the association between type of HCP initially contacted by an individual with NP, service utilization, and total episode cost. DCs were the most common initial contact HCP and were associated with the lowest adjusted total episode cost. The rate and timing of use of different services was highly variable among types of HCPs initially contacted by an individual with NP. Demographics of individuals varied by type of HCP initially contacted. The findings were consistent for individuals experiencing single or multiple episodes during the study period.

There are several limitations to consider when analyzing the observed associations. Data errors, missing information, and variability in benefit plan design and enrollee cost-sharing responsibility were potential sources of confounding or bias. These are partially addressed through extensive quality and actuarial control measures applied to source data. The commercial insurer HCP database, while under continual validation, may have included errors or missing information. Summarizing total episode cost has several potential limitations associated with insurance coverage, nature of network participation, and alternative reimbursement models. Individuals were from all 50 states and most US territories; however, this was not a U.S representative sample.

There are tradeoffs and potential limitations associated with the episode of care unit of analysis compared with a static, duration-based unit of analysis. For the 17.2% of individuals with multiple episodes during the study period, the study does not provide a summary of an individual’s experience across all episodes. The episode sequence cohort analyses reveal homogeneity in both initial contact HCP and subsequent care delivery whether NP is a single episode event, or a series of sequential episodes. This reduces the risk of severity confounding. The episode of care unit of measurement has potential translational benefits supporting the transition from fee for service to value-based episodic bundled payment arrangements.

There were potentially meaningful, yet unknown, differences in the; clinical complexity of individuals seeking treatment from different types of HCPs, preferences for the type of initial contact HCP, and expectations or requests for specific health care services. These limitations were partially addressed by excluding diagnoses involving complex pathology, performing a separate analysis for a non-surgical sample, performing a separate analysis for each episode sequence cohort, and utilizing a mixed effects models that included individual demographic variables, comorbidities using an ERG^®^ score, and a random effect to address variation in decision-making among individual HCPs of the same type.

Social disadvantage, population race/ethnicity, and geographic variation in HCP availability are important factors associated with variability in the treatment of NP and represent a translational limitation. In this study, the availability of non-prescribing HCPs had an inverse association with both the ADI and percent non-white population residing in a ZIP code. Less availability of non-prescribing HCPs may be associated with higher rates of prescription medication use, including opioids, for NP. These findings were beyond the scope of this paper are the focus of a subsequent study.

The analysis corroborated and expanded upon the findings of earlier work demonstrating the importance of the timing of access to non-prescribing HCPs for treatment of spinal disorders. This NP study used the same administrative data and methods as a previous LBP study^32^, and when considered together, provide a comprehensive analysis of the management of spinal disorders. Similar to LBP^26,32^ and corroborating a previous NP study^28^, when a non-prescribing HCP is the initial contact for NP, the rate of opioid use was found to be lower than with other types of initial contact HCPs. A potentially novel finding of this NP and the previous LBP study^32^ was the similarity with which each type of initial contact HCP approaches the management of NP and LBP (Supplement – Care Pathways). Fully exploring this similarity in approach to NP and LBP was beyond the scope of this paper and will be addressed in a subsequent paper.

This study corroborates previous work identifying the structure of the health care delivery system as a barrier to the delivery of guideline-concordant management of LBP and NP.^44^ Similar to LBP^32^, for NP two actions appear potentially important to improve guideline concordance and reduce cost of care. The first is supporting individual decision-making regarding the most appropriate type of initial HCP. With the persistent high rate of pharmaceutical, imaging, and interventional services identified in this study, this remains an important are of focus.^45,46^ The second is, when appropriate, supporting primary care and specialist HCPs in making timely referrals for non-pharmaceutical services. Potential barriers to greater use of non-pharmaceutical therapies identified in previous studies included the cost of non-pharmaceutical therapies, weak evidence supporting efficacy of non-pharmaceutical therapies, and administrative burden associated with referring individuals to a non-prescribing HCP.^47,48^

## Conclusions

Like LBP, the initial HCP selected by an individual with NP is associated with differences in both services received and total cost of care. Initial contact with a DC, LAc, PT, or DO providing spinal manipulation, was associated with an emphasis on non-pharmaceutical and non-interventional management. Initial contact with primary care, specialist, or emergency/urgent care HCPs was associated with an emphasis on pharmaceutical, imaging, and interventional services.

Compared to LBP, the relative absence of NP CPGs presents a barrier to understanding the degree to which observed variability is unwarranted. Like LBP, for individuals with NP, there appears to be an opportunity to identify strategies and tactics to make it easier for individuals to obtain non-pharmaceutical services through either selection of an initial contact HCP, or timely referral from primary care and specialist HCPs.

## Supporting information

Supplement - Care Pathway

Supplement - Cohort

Supplement - STROBE Checklist

Supplement 1 - Episodes By State

Supplement 2 - Episode Sequence Cohort

Supplement 3 - Non-surgical %

Supplement 3a - Non-surgical Risk Ratio

Supplement 4 - Pooled %

Supplement 4a - Pooled Risk Ratio

Supplement 5 - Non-surgical Timing

Supplement 6 - Pooled Timing

Supplement 7 - Pooled Cost

Supplement 8 - Pooled Mixed Effects Model

## Data Availability

All data produced in the present study are available upon reasonable request to the authors

## List of Abbreviations

NP: Neck pain
LBP: Low back pain
US: United States
YLD: Years lived with disability
DC: Doctor of Chiropractic
PT: Physical Therapist
EM: Emergency Medicine
HCP: Health care provider
LAc: Licensed Acupuncturist
ADI: Area Deprivation Index
STROBE: Strengthening the Reporting of Observational Studies in Epidemiology
ETG^*®*^: Episode Treatment Group^*®*^
ERG^*®*^: Episode Risk Group^*®*^
ACP: American College of Physicians
DO: Doctor of Osteopathy
OTC: Over the counter
PA: Physician’s Assistant
PCP: Primary care provider
OS: Orthopedic Surgeon
NS: Neurosurgeon
PMR: Physical Medicine and Rehabilitation
PM: Pain Management
UC: Urgent Care
Neuro: Neurologist
Rheum: Rheumatologist
CMT: Chiropractic manipulative treatment
OMT: Osteopathic manipulative treatment
OR: Odds ratio
RR: Risk ratio

## Declarations

### Ethics approval and consent to participate

The UnitedHealth Group Office of Human Research Affairs Institutional Review Board

### Consent for publication

Not applicable

### Availability of data and materials

The data are proprietary and are not available for public use but, under certain conditions, may be made available to editors and their approved auditors under a data-use agreement to confirm the findings of the current study.

### Authors’ contributions

Study conception and design: **DE**. Data acquisition: **DE, MZ**. Data analysis and interpretation: **DE, MZ**. Draft or substantially revise manuscript: **DE, MZ**.

## Acknowledgements

Amy Okaya and Elaine Preimesberger edited and contributed editorial recommendations that improved the manuscript. Thomas Kosloff, DC, provided important input on the study topic, methods, and discussion.

