## Supplement - Care Pathway for "Neck pain care pathways and costs: association with the type of initial contact health care provider. A retrospective cohort study"

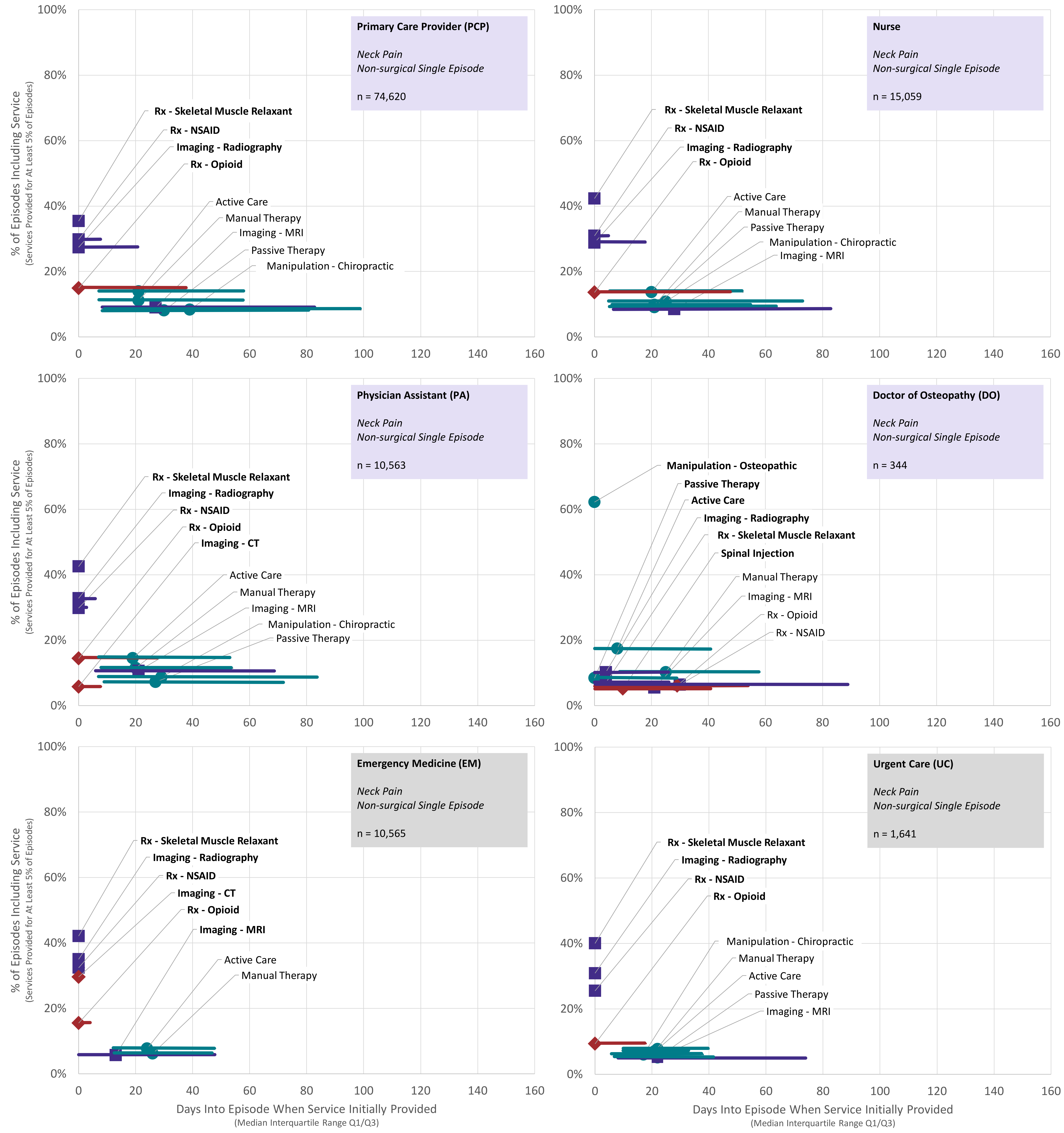


**Figure 2 Supplement.** Rate and timing of use of health care services for individuals with neck pain initially contacting a primary care provider, nurse, physician assistant, doctor of osteopathy, emergency medicine, and urgent care health care providers - non-surgical single episode cohort

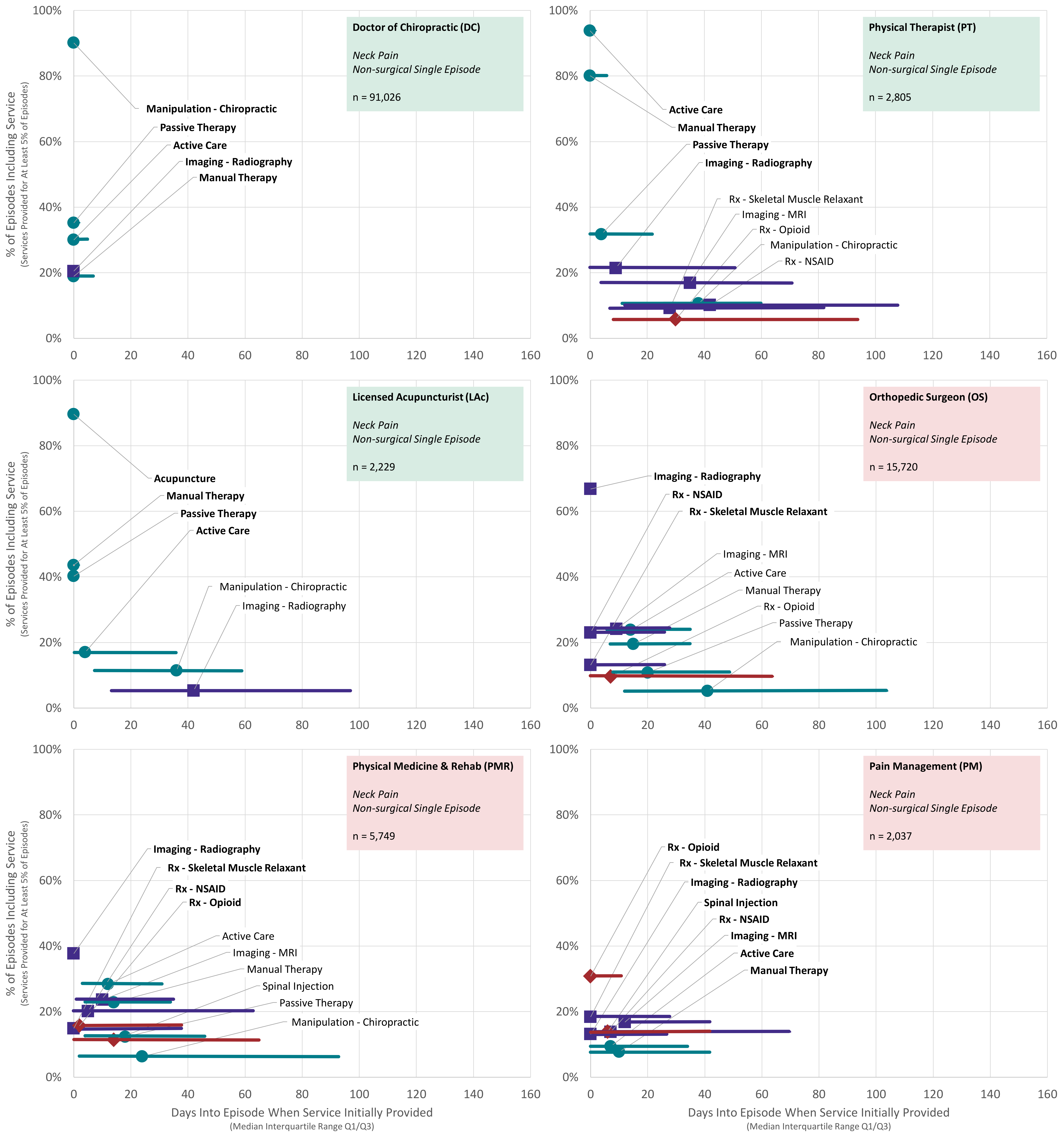


**Figure 2 Supplement.** Rate and timing of use of health care services for individuals with neck pain initially contacting a doctor of chiropractic, physical therapist, licensed acupuncturist assistant, orthopedic surgeon, physical medicine & rehabilitation, and pain management health care providers - non-surgical single episode cohort
