## Supplement 1 - Episodes By State for "Neck pain care pathways and costs: association with the type of initial contact health care provider. A retrospective cohort study"

### Supplement 1 - Neck pain episodes by individual home address state

| State | Pooled | | Non Surgical | | % Surgical |
|---|---|---|---|---|---|
| | Episodes | % of Total | Episodes | % of Total | |
| Total | 390992 | 100.0% | 374403 | 100.0% | 4.2% |
| TX | 44610 | 11.4% | 42317 | 11.3% | 5.1% |
| FL | 37375 | 9.6% | 35834 | 9.6% | 4.1% |
| MO | 19158 | 4.9% | 18448 | 4.9% | 3.7% |
| CA | 18524 | 4.7% | 17993 | 4.8% | 2.9% |
| OH | 17907 | 4.6% | 17267 | 4.6% | 3.6% |
| IL | 17398 | 4.5% | 16698 | 4.5% | 4.0% |
| WI | 17205 | 4.4% | 16528 | 4.4% | 3.9% |
| MN | 15435 | 4.0% | 14668 | 3.9% | 5.0% |
| NC | 14143 | 3.6% | 13529 | 3.6% | 4.3% |
| CO | 13852 | 3.5% | 13171 | 3.5% | 4.9% |
| AZ | 12574 | 3.2% | 11989 | 3.2% | 4.7% |
| GA | 11884 | 3.0% | 11332 | 3.0% | 4.6% |
| NY | 11300 | 2.9% | 10885 | 2.9% | 3.7% |
| MD | 10444 | 2.7% | 9965 | 2.7% | 4.6% |
| VA | 10418 | 2.7% | 9946 | 2.7% | 4.5% |
| IN | 8325 | 2.1% | 7898 | 2.1% | 5.1% |
| LA | 7917 | 2.0% | 7430 | 2.0% | 6.2% |
| IA | 7612 | 2.0% | 7407 | 2.0% | 2.7% |
| NE | 7562 | 1.9% | 7320 | 2.0% | 3.2% |
| TN | 7334 | 1.9% | 6997 | 1.9% | 4.6% |
| PA | 7104 | 1.8% | 6820 | 1.8% | 4.0% |
| WA | 6446 | 1.7% | 6296 | 1.7% | 2.3% |
| NJ | 5804 | 1.5% | 5612 | 1.5% | 3.3% |
| OK | 5104 | 1.3% | 4852 | 1.3% | 4.9% |
| AR | 4410 | 1.1% | 4191 | 1.1% | 5.0% |
| MI | 4363 | 1.1% | 4221 | 1.1% | 3.3% |
| KS | 4308 | 1.1% | 4090 | 1.1% | 5.1% |

| State | Pooled | | Non Surgical | | % Surgical |
|---|---|---|---|---|---|
| | Episodes | % of Total | Episodes | % of Total | |
| OR | 3935 | 1.0% | 3848 | 1.0% | 2.2% |
| MS | 3928 | 1.0% | 3671 | 1.0% | 6.5% |
| KY | 3702 | 1.0% | 3555 | 1.0% | 4.0% |
| CT | 3401 | 0.9% | 3286 | 0.9% | 3.4% |
| UT | 3361 | 0.9% | 3173 | 0.9% | 5.6% |
| MA | 3283 | 0.8% | 3174 | 0.9% | 3.3% |
| SC | 3199 | 0.8% | 2994 | 0.8% | 6.4% |
| RI | 2670 | 0.7% | 2596 | 0.7% | 2.8% |
| NV | 1985 | 0.5% | 1896 | 0.5% | 4.5% |
| AL | 1903 | 0.5% | 1780 | 0.5% | 6.5% |
| NM | 1296 | 0.3% | 1269 | 0.3% | 2.1% |
| DC | 1102 | 0.3% | 1076 | 0.3% | 2.4% |
| ND | 1097 | 0.3% | 1067 | 0.3% | 2.7% |
| WV | 703 | 0.2% | 680 | 0.2% | 3.3% |
| NH | 673 | 0.2% | 646 | 0.2% | 4.0% |
| SD | 576 | 0.2% | 560 | 0.2% | 2.8% |
| ID | 554 | 0.1% | 535 | 0.1% | 3.4% |
| ME | 465 | 0.1% | 450 | 0.1% | 3.2% |
| DE | 357 | 0.1% | 341 | 0.1% | 4.5% |
| WY | 343 | 0.1% | 321 | 0.1% | 6.4% |
| VI | 337 | 0.1% | 333 | 0.1% | 1.2% |
| MT | 244 | 0.1% | 235 | 0.1% | 3.7% |
| VT | 86 | 0.0% | 85 | 0.0% | 1.2% |
| AK | 62 | 0.0% | 55 | 0.0% | 11.3% |
| HI | 55 | 0.0% | 52 | 0.0% | 5.5% |
| PR | 49 | 0.0% | 48 | 0.0% | 2.0% |
| Unknown | 3110 | 0.8% | 2973 | 0.8% | 4.4% |

Pooled = Combined surgical and non-surgical episodes
