## Supplement 2 - Episode Sequence Cohort for "Neck pain care pathways and costs: association with the type of initial contact health care provider. A retrospective cohort study"

### Supplement 2 - Neck pain episode sequence cohorts

| Individuals | | | Episode Sequence Cohorts | | | | Time Intervals (days) - Median (IQR Q1,Q3)(Minimum) | | | |
|---|---|---|---|---|---|---|---|---|---|---|
| | Count | % | | | Episodes | % | Episode Duration | Clean Period - Before Initial Episode | Clean Period - Between Sequential Episodes | Clean Period - After Final Episode |
| 1 Episode | 267615 | 82.8% | Single Episode | | 267615 | 68.5% | 24 (1, 117) (1) | 650 (438, 865) (91) | N/A | 417 (251, 655) (61) |
| 2 Episodes | 46077 | 14.2% | Multiple Episodes | First | 55733 | 14.3% | 55 (3, 178) (1) | 373 (212, 558) (91) | N/A | N/A |
| 3 Episodes | 7995 | 2.5% | | Second | 55733 | 14.3% | 32 (1, 136) (1) | N/A | 198 (119, 332) (61) | 383 (209, 530) (61) |
| 4+ Episodes | 1661 | 0.5% | | Third+ | 11911 | 3.1% | 11 (1, 78) (1) | N/A | 161 (108, 264) (61) | 230 (134, 363) (61) |
| Total | 323348 | 100.0% | Total | | 390992 | 100.0% | 29 (1, 128) (1) | 597 (370, 831) (91) | 189 (116, 316) (61) | 401 (236, 617) (61) |
