## Supplement 3 - Non-surgical % for "Neck pain care pathways and costs: association with the type of initial contact health care provider. A retrospective cohort study"

| Supplement 3 - Neck pain % of episodes including service by type of initial contact health care provider (HCP) | | | | | | | | | | | | | | | | | | | | |
|---|---|---|---|---|---|---|---|---|---|---|---|---|---|---|---|---|---|---|---|---|
| All Non-Surgical Episodes | | Episodes | | First Line | | | | | | | Second Line | | | | | Third Line | | | | |
| | | Count | % | *Any* | Manipulation - Chiropractic | Active Care | Passive Therapy | Manual Therapy | Acupuncture | Manipulation - Osteopathic | *Any* | Imaging - Radiography | Rx - NSAID | Rx - Skeletal Muscle Relaxant | Imaging - MRI | *Any* | Rx-Opioid | Spinal Injection | Spinal Surgery | Imaging-CT |
| Total | | *374403* | *100.0%* | *52.4%* | *41.4%* | *20.8%* | *18.9%* | *15.3%* | *1.3%* | *1.2%* | *48.9%* | *25.0%* | *17.6%* | *18.4%* | *8.7%* | *14.8%* | *9.9%* | *2.5%* | *0.0%* | *3.7%* |
| Primary Care | PCP | 103095 | 25.8% | *22.6%* | 9.0% | 13.6% | 8.2% | 11.0% | 0.4% | 2.8% | *65.2%* | 25.1% | 30.3% | 33.2% | 9.2% | *19.9%* | 15.9% | 2.6% | 0.0% | 3.0% |
| | Nurse | 20287 | 5.1% | *21.9%* | 11.4% | 13.5% | 8.9% | 9.7% | 0.3% | 0.7% | *70.7%* | 26.3% | 31.3% | 39.8% | 8.7% | *21.2%* | 14.8% | 4.4% | 0.0% | 4.1% |
| | PA | 13892 | 3.5% | *21.3%* | 9.6% | 14.4% | 7.5% | 11.4% | 0.3% | 0.4% | *74.6%* | 30.8% | 30.7% | 40.0% | 11.1% | *22.6%* | 15.5% | 4.0% | 0.0% | 5.3% |
| | DO | 522 | 0.1% | *69.0%* | 3.1% | 14.6% | 7.1% | 10.0% | 0.6% | 63.8% | *23.2%* | 9.4% | 6.9% | 8.4% | 5.0% | *10.9%* | 5.7% | 5.2% | 0.0% | 1.0% |
| | *All* | *137796* | *34.5%* | *22.5%* | *9.4%* | *13.7%* | *8.2%* | *10.8%* | *0.4%* | *2.5%* | *66.8%* | *25.8%* | *30.4%* | *34.7%* | *9.3%* | *20.3%* | *15.6%* | *3.0%* | *0.0%* | *3.4%* |
| Non-Prescriber | DC | 149092 | 37.3% | *95.6%* | 91.7% | 28.6% | 34.0% | 18.7% | 0.7% | 0.1% | *21.8%* | 16.1% | 4.5% | 3.6% | 1.9% | *4.3%* | 3.2% | 0.8% | 0.0% | 0.6% |
| | PT | 3813 | 1.0% | *97.4%* | 11.7% | 93.6% | 32.8% | 80.7% | 1.9% | 0.8% | *40.2%* | 21.2% | 10.9% | 9.4% | 16.6% | *11.8%* | 6.2% | 3.9% | 0.0% | 2.9% |
| | LAc | 3191 | 0.8% | *97.3%* | 12.4% | 18.2% | 41.0% | 45.0% | 88.3% | 0.9% | *10.2%* | 5.0% | 3.4% | 2.5% | 2.9% | *4.5%* | 3.2% | 1.4% | 0.0% | 0.3% |
| | *All* | *156096* | *39.0%* | *95.7%* | *88.1%* | *30.0%* | *34.1%* | *20.8%* | *2.6%* | *0.2%* | *22.0%* | *16.0%* | *4.6%* | *3.7%* | *2.3%* | *4.5%* | *3.3%* | *0.9%* | *0.0%* | *0.6%* |
| Specialist | OS | 21266 | 5.3% | *27.2%* | 5.5% | 23.2% | 10.7% | 19.0% | 0.4% | 0.6% | *83.1%* | 63.5% | 24.6% | 13.1% | 24.1% | *16.5%* | 10.6% | 5.0% | 0.0% | 2.8% |
| | PMR | 8306 | 2.1% | *33.1%* | 6.2% | 27.7% | 12.6% | 22.4% | 1.8% | 2.5% | *62.7%* | 33.7% | 20.0% | 15.3% | 22.7% | *27.5%* | 17.0% | 12.0% | 0.0% | 2.4% |
| | PM | 3161 | 0.8% | *10.7%* | 2.0% | 8.5% | 3.3% | 6.9% | 0.2% | 0.6% | *43.8%* | 11.6% | 13.9% | 17.0% | 14.9% | *41.9%* | 30.8% | 14.2% | 0.0% | 2.0% |
| | NS | 3798 | 0.9% | *20.7%* | 4.0% | 17.5% | 8.3% | 14.5% | 0.3% | 0.2% | *75.1%* | 41.9% | 17.0% | 15.4% | 41.4% | *23.3%* | 14.2% | 5.0% | 0.0% | 8.3% |
| | Neuro | 7950 | 2.0% | *19.0%* | 5.9% | 14.6% | 7.6% | 11.7% | 0.4% | 0.5% | *63.4%* | 12.4% | 19.4% | 27.3% | 28.6% | *20.2%* | 11.1% | 7.8% | 0.0% | 3.6% |
| | Rheu | 3305 | 0.8% | *17.8%* | 8.3% | 12.0% | 7.0% | 9.7% | 0.5% | 0.4% | *69.6%* | 33.1% | 26.9% | 25.1% | 12.6% | *21.6%* | 15.0% | 6.3% | 0.0% | 2.6% |
| | MD (Oth) | 7239 | 1.8% | *24.3%* | 14.2% | 12.2% | 8.7% | 9.4% | 0.7% | 2.5% | *43.7%* | 17.1% | 20.8% | 15.7% | 9.0% | *27.5%* | 22.2% | 2.5% | 0.0% | 4.7% |
| | *All* | *55025* | *13.8%* | *24.6%* | *6.7%* | *19.3%* | *9.5%* | *15.7%* | *0.6%* | *1.1%* | *68.4%* | *39.2%* | *21.7%* | *16.9%* | *22.5%* | *22.4%* | *14.8%* | *6.8%* | *0.0%* | *3.5%* |
| Emergency/ Urgent Care | EM | 12124 | 3.0% | *12.1%* | 5.4% | 8.3% | 5.1% | 6.7% | 0.2% | 0.3% | *70.9%* | 34.6% | 33.1% | 41.4% | 6.4% | *41.4%* | 16.3% | 1.4% | 0.0% | 28.9% |
| | Rad | 11430 | 2.9% | *4.3%* | 0.8% | 3.6% | 1.5% | 2.9% | 0.1% | 0.0% | *80.4%* | 58.6% | 1.6% | 1.5% | 26.9% | *24.5%* | 1.0% | 0.9% | 0.0% | 23.0% |
| | UC | 1932 | 0.5% | *14.4%* | 6.6% | 8.2% | 5.5% | 7.8% | 0.3% | 1.2% | *65.6%* | 30.0% | 26.7% | 38.9% | 6.0% | *13.2%* | 9.9% | 1.4% | 0.0% | 3.1% |
| | *All* | *25486* | *6.4%* | *8.8%* | *3.4%* | *6.2%* | *3.5%* | *5.1%* | *0.2%* | *0.2%* | *74.8%* | *45.0%* | *18.5%* | *23.3%* | *15.6%* | *31.7%* | *8.9%* | *1.2%* | *0.0%* | *24.3%* |

PCP=Primary Care Provider, PA=Physician Assistant, DO=Doctor of Osteopathy, DC=Doctor of Chiropractic, PT=Physical Therapist, LAc=Licensed Acupuncturist, OS=Orthpedic Surgeon, PMR=Physical Medicine & Rehabilitation, PM=Pain Management, NS=Neurosurgeon, Neuro=Neurologist, Rheum=Rheumatologist, MD Oth=Other MD specialty, EM=Emergency Medicine, Rad=Radiologist, UC=Urgent Care
