## Supplement 3a - Non-surgical Risk Ratio for "Neck pain care pathways and costs: association with the type of initial contact health care provider. A retrospective cohort study"

**Supplement 3a - Risk ratio and 95% confidence interval for service use during non-surgical neck pain episodes by type of initial contact health care provider as compared to a primary care provider (PCP)**

| All Non-Surgical Episodes | | First Line | | | | | | | Second Line | | | | | Third Line | | | | |
|---|---|---|---|---|---|---|---|---|---|---|---|---|---|---|---|---|---|---|
| | | Any | Manipulation - Chiropractic | Active Care | Passive Therapy | Manual Therapy | Acupuncture | Manipulation - Osteopathic | Any | Imaging - Radiography | Rx - NSAID | Rx - Skeletal Muscle Relaxant | Imaging - MRI | Any | Rx-Opioid | Spinal Injection | Spinal Surgery | Imaging-CT |
| Primary Care | PCP | reference | | | | | | | | | | | | | | | | |
| Primary Care | Nurse | 0.97 (0.95, 1.00) | 1.26 (1.20, 1.31) | 0.99 (0.95, 1.03) | 1.08 (1.03, 1.14) | 0.89 (0.85, 0.93) | 0.68 (0.52, 0.89) | 0.24 (0.21, 0.29) | 1.08 (1.07, 1.10) | 1.05 (1.02, 1.08) | 1.03 (1.01, 1.06) | 1.20 (1.18, 1.22) | 0.95 (0.90, 0.99) | 1.07 (1.04, 1.10) | 0.93 (0.90, 0.97) | 1.69 (1.57, 1.83) | N/A | 1.36 (1.27, 1.47) |
| Primary Care | PA | 0.94 (0.91, 0.98) | 1.06 (1.00, 1.12) | 1.06 (1.02, 1.11) | 0.91 (0.85, 0.97) | 1.04 (0.99, 1.09) | 0.65 (0.47, 0.90) | 0.15 (0.12, 0.19) | 1.15 (1.13, 1.16) | 1.23 (1.20, 1.26) | 1.01 (0.99, 1.04) | 1.21 (1.18, 1.23) | 1.21 (1.15, 1.27) | 1.14 (1.10, 1.17) | 0.98 (0.94, 1.02) | 1.55 (1.41, 1.69) | N/A | 1.75 (1.62, 1.89) |
| Primary Care | DO | 3.06 (2.88, 3.24) | 0.34 (0.21, 0.55) | 1.07 (0.87, 1.32) | 0.86 (0.63, 1.18) | 0.91 (0.70, 1.18) | 1.30 (0.42, 4.02) | 22.53 (20.93, 24.26) | 0.36 (0.30, 0.42) | 0.37 (0.29, 0.49) | 0.23 (0.17, 0.31) | 0.25 (0.19, 0.34) | 0.54 (0.37, 0.79) | 0.55 (0.43, 0.70) | 0.36 (0.26, 0.51) | 2.01 (1.39, 2.91) | N/A | 0.32 (0.13, 0.76) |
| Non-Prescriber | DC | 4.24 (4.19, 4.28) | 10.14 (9.95, 10.34) | 2.10 (2.06, 2.14) | 4.13 (4.04, 4.22) | 1.71 (1.67, 1.74) | 1.66 (1.49, 1.86) | 0.05 (0.04, 0.06) | 0.33 (0.33, 0.34) | 0.64 (0.63, 0.65) | 0.15 (0.15, 0.15) | 0.11 (0.10, 0.11) | 0.21 (0.20, 0.22) | 0.22 (0.21, 0.22) | 0.20 (0.19, 0.21) | 0.30 (0.28, 0.32) | N/A | 0.19 (0.18, 0.21) |
| Non-Prescriber | PT | 4.31 (4.26, 4.37) | 1.29 (1.18, 1.41) | 6.87 (6.75, 7.00) | 3.99 (3.80, 4.20) | 7.36 (7.19, 7.53) | 4.38 (3.43, 5.58) | 0.27 (0.19, 0.39) | 0.62 (0.59, 0.64) | 0.84 (0.79, 0.90) | 0.36 (0.33, 0.40) | 0.28 (0.26, 0.31) | 1.81 (1.68, 1.95) | 0.59 (0.54, 0.65) | 0.39 (0.35, 0.45) | 1.53 (1.30, 1.80) | N/A | 0.97 (0.80, 1.16) |
| Non-Prescriber | LAc | 4.31 (4.26, 4.37) | 1.38 (1.25, 1.51) | 1.34 (1.24, 1.44) | 4.99 (4.76, 5.23) | 4.11 (3.94, 4.28) | 199.15 (181.58, 218.42) | 0.33 (0.23, 0.47) | 0.16 (0.14, 0.17) | 0.20 (0.17, 0.23) | 0.11 (0.09, 0.14) | 0.08 (0.06, 0.09) | 0.31 (0.25, 0.38) | 0.23 (0.19, 0.26) | 0.20 (0.16, 0.24) | 0.54 (0.40, 0.72) | N/A | 0.11 (0.06, 0.21) |
| Specialist | OS | 1.20 (1.18, 1.24) | 0.61 (0.58, 0.65) | 1.70 (1.65, 1.75) | 1.30 (1.24, 1.36) | 1.74 (1.68, 1.79) | 0.81 (0.63, 1.03) | 0.22 (0.18, 0.26) | 1.28 (1.27, 1.29) | 2.54 (2.50, 2.57) | 0.81 (0.79, 0.83) | 0.39 (0.38, 0.41) | 2.63 (2.55, 2.71) | 0.83 (0.80, 0.86) | 0.67 (0.64, 0.70) | 1.95 (1.82, 2.09) | N/A | 0.94 (0.87, 1.03) |
| Specialist | PMR | 1.47 (1.42, 1.52) | 0.69 (0.63, 0.75) | 2.03 (1.96, 2.11) | 1.54 (1.45, 1.63) | 2.04 (1.95, 2.13) | 4.07 (3.39, 4.89) | 0.90 (0.78, 1.03) | 0.96 (0.95, 0.98) | 1.35 (1.30, 1.39) | 0.66 (0.63, 0.69) | 0.46 (0.44, 0.49) | 2.47 (2.37, 2.59) | 1.38 (1.33, 1.44) | 1.07 (1.02, 1.12) | 4.68 (4.37, 5.02) | N/A | 0.80 (0.69, 0.92) |
| Specialist | PM | 0.47 (0.43, 0.52) | 0.22 (0.18, 0.29) | 0.63 (0.56, 0.70) | 0.40 (0.33, 0.49) | 0.63 (0.56, 0.72) | 0.50 (0.24, 1.05) | 0.22 (0.14, 0.35) | 0.67 (0.65, 0.70) | 0.46 (0.42, 0.51) | 0.46 (0.42, 0.50) | 0.51 (0.47, 0.55) | 1.62 (1.49, 1.76) | 2.11 (2.02, 2.20) | 1.94 (1.84, 2.05) | 5.54 (5.04, 6.08) | N/A | 0.66 (0.52, 0.85) |
| Specialist | Neuro | 0.92 (0.86, 0.98) | 0.44 (0.38, 0.52) | 1.28 (1.19, 1.38) | 1.01 (0.91, 1.12) | 1.32 (1.22, 1.43) | 0.59 (0.32, 1.11) | 0.08 (0.04, 0.16) | 1.15 (1.13, 1.17) | 1.67 (1.61, 1.74) | 0.56 (0.52, 0.60) | 0.47 (0.43, 0.50) | 4.51 (4.32, 4.70) | 1.17 (1.11, 1.24) | 0.90 (0.83, 0.97) | 1.94 (1.68, 2.23) | N/A | 2.77 (2.48, 3.10) |
| Specialist | Neurologist | 0.84 (0.80, 0.88) | 0.65 (0.59, 0.71) | 1.07 (1.02, 1.13) | 0.92 (0.85, 1.00) | 1.07 (1.00, 1.14) | 0.94 (0.66, 1.33) | 0.16 (0.12, 0.23) | 0.97 (0.96, 0.99) | 0.50 (0.47, 0.53) | 0.64 (0.61, 0.67) | 0.82 (0.79, 0.86) | 3.12 (3.00, 3.24) | 1.01 (0.97, 1.06) | 0.70 (0.66, 0.75) | 3.04 (2.80, 3.31) | N/A | 1.21 (1.08, 1.36) |
| Specialist | Rheu | 0.79 (0.73, 0.85) | 0.92 (0.82, 1.03) | 0.88 (0.80, 0.96) | 0.85 (0.75, 0.96) | 0.88 (0.79, 0.98) | 1.09 (0.66, 1.80) | 0.14 (0.08, 0.24) | 1.07 (1.04, 1.09) | 1.32 (1.26, 1.39) | 0.89 (0.84, 0.94) | 0.76 (0.71, 0.80) | 1.37 (1.25, 1.51) | 1.09 (1.02, 1.16) | 0.95 (0.87, 1.03) | 2.46 (2.15, 2.82) | N/A | 0.86 (0.70, 1.07) |
| Specialist | MD (Oth) | 1.08 (1.03, 1.12) | 1.57 (1.48, 1.67) | 0.90 (0.84, 0.96) | 1.06 (0.98, 1.14) | 0.86 (0.80, 0.93) | 1.65 (1.24, 2.19) | 0.89 (0.77, 1.03) | 0.67 (0.65, 0.69) | 0.68 (0.65, 0.72) | 0.69 (0.66, 0.72) | 0.47 (0.45, 0.50) | 0.98 (0.91, 1.06) | 1.38 (1.33, 1.44) | 1.40 (1.34, 1.46) | 0.97 (0.83, 1.12) | N/A | 1.57 (1.41, 1.75) |
| Emergency /Urgent Care | EM | 0.54 (0.51, 0.56) | 0.59 (0.55, 0.64) | 0.61 (0.57, 0.65) | 0.62 (0.58, 0.67) | 0.61 (0.57, 0.65) | 0.54 (0.37, 0.79) | 0.09 (0.07, 0.13) | 1.09 (1.07, 1.10) | 1.38 (1.34, 1.42) | 1.09 (1.06, 1.12) | 1.25 (1.22, 1.28) | 0.69 (0.64, 0.74) | 2.08 (2.03, 2.13) | 1.03 (0.98, 1.07) | 0.55 (0.47, 0.64) | N/A | 9.59 (9.17, 10.02) |
| Emergency /Urgent Care | Rad | 0.19 (0.17, 0.21) | 0.08 (0.07, 0.10) | 0.27 (0.24, 0.29) | 0.19 (0.16, 0.22) | 0.27 (0.24, 0.30) | 0.22 (0.12, 0.39) | 0.02 (0.01, 0.04) | 1.23 (1.22, 1.25) | 2.34 (2.30, 2.38) | 0.05 (0.05, 0.06) | 0.05 (0.04, 0.05) | 2.93 (2.83, 3.04) | 1.23 (1.19, 1.27) | 0.06 (0.05, 0.08) | 0.33 (0.27, 0.41) | N/A | 7.65 (7.29, 8.03) |
| Emergency /Urgent Care | UC | 0.64 (0.57, 0.71) | 0.73 (0.62, 0.87) | 0.60 (0.52, 0.70) | 0.67 (0.55, 0.80) | 0.71 (0.61, 0.83) | 0.58 (0.24, 1.41) | 0.44 (0.29, 0.65) | 1.01 (0.97, 1.04) | 1.20 (1.12, 1.28) | 0.88 (0.82, 0.95) | 1.17 (1.11, 1.24) | 0.65 (0.54, 0.77) | 0.66 (0.59, 0.74) | 0.63 (0.55, 0.72) | 0.56 (0.39, 0.82) | N/A | 1.01 (0.79, 1.31) |

PCP=Primary Care Provider, PA=Physician Assistant, DO=Doctor of Osteopathy, DC=Doctor of Chiropractic, PT=Physical Therapist, LAc=Licensed Acupuncturist, OS=Orthpedic Surgeon, PMR=Physical Medicine & Rehabilitation, PM=Pain Management, NS=Neurosurgeon, Neuro=Neurologist, Rheum=Rheumatologist, MD Oth=Other MD specialty, EM=Emergency Medicine, Rad=Radiologist, UC=Urgent Care

Cells in red are not different than the PCP reference (p=.05)
