## Supplement 4 - Pooled % for "Neck pain care pathways and costs: association with the type of initial contact health care provider. A retrospective cohort study"

**Supplement 4 - Neck pain % of episodes including service by type of initial contact health care provider (HCP)**

| All Combined Surgical and Non-Surgical (Pooled) Episodes | | Episodes | | First Line | | | | | | | Second Line | | | | | Third Line | | | | |
|---|---|---|---|---|---|---|---|---|---|---|---|---|---|---|---|---|---|---|---|---|
| | | Count | % | *Any* | Manipulation - Chiropractic | Active Care | Passive Therapy | Manual Therapy | Acupuncture | Manipulation - Osteopathic | *Any* | Imaging - Radiography | Rx - NSAID | Rx - Skeletal Muscle Relaxant | Imaging - MRI | *Any* | Rx-Opioid | Spinal Injection | Spinal Surgery | Imaging-CT |
| *Total* | | *390992* | *100.0%* | *52.0%* | *40.2%* | *21.5%* | *18.9%* | *15.9%* | *1.3%* | *1.2%* | *50.5%* | *26.5%* | *18.3%* | *19.2%* | *11.0%* | *18.4%* | *11.6%* | *5.0%* | *4.2%* | *4.0%* |
| Primary Care | PCP | 107753 | 25.8% | *23.5%* | 9.1% | 14.6% | 8.7% | 11.8% | 0.5% | 2.8% | *66.3%* | 26.6% | 31.0% | 33.7% | 11.7% | *23.4%* | 17.6% | 5.1% | 4.3% | 3.4% |
| | Nurse | 21391 | 5.1% | *22.6%* | 11.2% | 14.3% | 9.2% | 10.4% | 0.3% | 0.7% | *71.4%* | 27.7% | 31.7% | 40.1% | 11.4% | *25.3%* | 16.6% | 7.6% | 5.2% | 4.5% |
| | PA | 14787 | 3.5% | *22.6%* | 9.6% | 15.8% | 8.1% | 12.5% | 0.3% | 0.4% | *75.7%* | 32.8% | 31.3% | 40.4% | 14.6% | *27.3%* | 17.6% | 7.6% | 6.1% | 5.5% |
| | DO | 533 | 0.1% | *69.2%* | 3.6% | 15.2% | 7.3% | 10.5% | 0.8% | 63.8% | *24.6%* | 10.3% | 7.1% | 8.6% | 6.2% | *12.8%* | 6.9% | 6.8% | 2.1% | 1.7% |
| | *All* | *144464* | *34.6%* | *23.4%* | *9.4%* | *14.7%* | *8.7%* | *11.6%* | *0.4%* | *2.5%* | *67.9%* | *27.4%* | *31.1%* | *35.3%* | *11.9%* | *24.0%* | *17.4%* | *5.7%* | *4.6%* | *3.8%* |
| Non-Prescriber | DC | 150381 | 36.0% | *95.6%* | 91.7% | 28.8% | 34.1% | 18.9% | 0.7% | 0.1% | *22.4%* | 16.5% | 4.7% | 3.9% | 2.5% | *5.1%* | 3.5% | 1.3% | 0.9% | 0.6% |
| | PT | 4111 | 1.0% | *97.5%* | 11.7% | 93.8% | 33.9% | 81.1% | 1.9% | 0.7% | *43.5%* | 23.6% | 12.0% | 10.7% | 20.0% | *18.2%* | 8.8% | 8.8% | 7.2% | 3.3% |
| | LAc | 3216 | 0.8% | *97.3%* | 12.5% | 18.4% | 41.0% | 45.2% | 88.2% | 0.9% | *10.7%* | 5.4% | 3.6% | 2.6% | 3.2% | *5.2%* | 3.4% | 1.8% | 0.8% | 0.4% |
| | *All* | *157708* | *37.8%* | *95.7%* | *88.0%* | *30.3%* | *34.3%* | *21.1%* | *2.6%* | *0.2%* | *22.7%* | *16.4%* | *4.9%* | *4.0%* | *2.9%* | *5.5%* | *3.7%* | *1.5%* | *1.0%* | *0.7%* |
| Specialist | OS | 24080 | 5.8% | *29.1%* | 5.6% | 25.2% | 11.6% | 20.4% | 0.4% | 0.6% | *84.5%* | 65.5% | 25.4% | 15.1% | 29.5% | *26.2%* | 15.3% | 11.3% | 11.7% | 4.0% |
| | PMR | 9487 | 2.3% | *33.3%* | 6.3% | 27.9% | 12.8% | 22.5% | 1.7% | 2.4% | *64.4%* | 35.0% | 21.0% | 16.5% | 26.6% | *36.5%* | 19.7% | 20.3% | 12.4% | 3.2% |
| | PM | 4076 | 1.0% | *10.4%* | 1.9% | 8.5% | 3.3% | 6.9% | 0.2% | 0.6% | *44.5%* | 12.5% | 13.3% | 16.1% | 18.4% | *55.0%* | 29.6% | 31.6% | 22.4% | 2.3% |
| | NS | 5066 | 1.2% | *26.0%* | 4.0% | 23.0% | 9.9% | 18.2% | 0.2% | 0.2% | *80.1%* | 51.0% | 18.4% | 21.7% | 46.2% | *42.5%* | 25.6% | 12.8% | 25.0% | 11.4% |
| | Neuro | 8854 | 2.1% | *20.0%* | 5.8% | 15.7% | 7.8% | 12.4% | 0.4% | 0.4% | *63.6%* | 14.9% | 19.3% | 27.6% | 30.1% | *28.3%* | 13.8% | 10.7% | 10.2% | 4.4% |
| | Rheu | 3488 | 0.8% | *18.8%* | 8.4% | 13.0% | 7.5% | 10.4% | 0.5% | 0.4% | *70.4%* | 34.0% | 27.4% | 25.5% | 14.8% | *25.7%* | 16.9% | 9.6% | 5.2% | 3.0% |
| | MD (Oth) | 7594 | 1.8% | *24.8%* | 14.1% | 13.0% | 8.9% | 10.0% | 0.7% | 2.4% | *45.5%* | 18.9% | 21.4% | 16.8% | 11.0% | *30.9%* | 23.5% | 5.0% | 4.7% | 5.3% |
| | *All* | *62645* | *15.0%* | *25.9%* | *6.5%* | *20.8%* | *10.0%* | *16.7%* | *0.6%* | *1.0%* | *70.1%* | *41.7%* | *22.2%* | *18.5%* | *26.7%* | *31.8%* | *18.6%* | *13.2%* | *12.2%* | *4.5%* |
| Emergency/ Urgent Care | EM | 12479 | 3.0% | *13.0%* | 5.5% | 9.1% | 5.6% | 7.4% | 0.2% | 0.3% | *71.4%* | 35.5% | 33.3% | 41.7% | 8.0% | *43.0%* | 17.4% | 2.9% | 2.8% | 29.0% |
| | Rad | 11717 | 2.8% | *4.6%* | 0.8% | 4.0% | 1.7% | 3.2% | 0.1% | 0.1% | *80.5%* | 58.3% | 1.8% | 1.8% | 28.1% | *26.3%* | 1.5% | 2.5% | 2.4% | 22.9% |
| | UC | 1979 | 0.5% | *15.2%* | 6.7% | 9.0% | 5.8% | 8.4% | 0.3% | 1.2% | *66.2%* | 30.9% | 27.0% | 39.1% | 7.5% | *15.3%* | 10.8% | 2.8% | 2.4% | 3.2% |
| | *All* | *26175* | *6.3%* | *9.4%* | *3.5%* | *6.8%* | *3.9%* | *5.6%* | *0.2%* | *0.2%* | *75.1%* | *45.4%* | *18.8%* | *23.6%* | *17.0%* | *33.5%* | *9.8%* | *2.7%* | *2.6%* | *24.3%* |

PCP=Primary Care Provider, PA=Physician Assistant, DO=Doctor of Osteopathy, DC=Doctor of Chiropractic, PT=Physical Therapist, LAc=Licensed Acupuncturist, OS=Orthpedic Surgeon, PMR=Physical Medicine & Rehabilitation, PM=Pain Management, NS=Neurosurgeon, Neuro=Neurologist, Rheum=Rheumatologist, MD Oth=Other MD specialty, EM=Emergency Medicine, Rad=Radiologist, UC=Urgent Care
