## Supplement 4a - Pooled Risk Ratio for "Neck pain care pathways and costs: association with the type of initial contact health care provider. A retrospective cohort study"

**Supplement 4a - Risk ratio and 95% confidence interval for service use during combined surgical and non-surgical (pooled) neck pain episodes by type of initial contact health care provider as compared to a primary care provider (PCP)**

| Combined Surgical and Non-surgical (Pooled) Episodes | | First Line | | | | | | | Second Line | | | | | Third Line | | | | |
|---|---|---|---|---|---|---|---|---|---|---|---|---|---|---|---|---|---|---|
| | | Any | Manipulation - Chiropractic | Active Care | Passive Therapy | Manual Therapy | Acupuncture | Manipulation - Osteopathic | Any | Imaging - Radiography | Rx - NSAID | Rx - Skeletal Muscle Relaxant | Imaging - MRI | Any | Rx-Opioid | Spinal Injection | Spinal Surgery | Imaging-CT |
| Primary Care | PCP | reference | | | | | | | | | | | | | | | | |
| | Nurse | 0.96 (0.94, 0.99) | 1.24 (1.19, 1.29) | 0.98 (0.95, 1.02) | 1.06 (1.01, 1.11) | 0.88 (0.84, 0.92) | 0.67 (0.52, 0.87) | 0.25 (0.21, 0.30) | 1.08 (1.07, 1.09) | 1.04 (1.01, 1.06) | 1.02 (1.00, 1.05) | 1.19 (1.17, 1.21) | 0.97 (0.93, 1.01) | 1.08 (1.05, 1.11) | 0.94 (0.91, 0.97) | 1.48 (1.40, 1.56) | 1.19 (1.12, 1.27) | 1.31 (1.22, 1.41) |
| | PA | 0.96 (0.93, 0.99) | 1.05 (1.00, 1.11) | 1.08 (1.04, 1.13) | 0.93 (0.88, 0.98) | 1.06 (1.02, 1.11) | 0.73 (0.54, 0.98) | 0.16 (0.12, 0.20) | 1.14 (1.13, 1.15) | 1.23 (1.20, 1.26) | 1.01 (0.98, 1.03) | 1.20 (1.17, 1.22) | 1.24 (1.19, 1.30) | 1.17 (1.14, 1.20) | 1.00 (0.96, 1.04) | 1.48 (1.39, 1.57) | 1.40 (1.31, 1.50) | 1.63 (1.51, 1.75) |
| | DO | 2.95 (2.79, 3.13) | 0.39 (0.25, 0.61) | 1.04 (0.85, 1.27) | 0.84 (0.62, 1.14) | 0.89 (0.70, 1.14) | 1.65 (0.62, 4.40) | 23.01 (21.39, | 0.37 (0.32, 0.43) | 0.39 (0.30, 0.50) | 0.23 (0.17, 0.31) | 0.26 (0.19, 0.34) | 0.53 (0.38, 0.74) | 0.55 (0.44, 0.68) | 0.39 (0.29, 0.54) | 1.32 (0.96, 1.81) | 0.48 (0.27, 0.86) | 0.50 (0.26, 0.95) |
| Non-Prescriber | DC | 4.08 (4.03, 4.12) | 10.10 (9.91, 10.29) | 1.97 (1.94, 2.01) | 3.92 (3.84, 4.00) | 1.61 (1.58, 1.64) | 1.64 (1.48, 1.83) | 0.05 (0.05, 0.06) | 0.34 (0.33, 0.34) | 0.62 (0.61, 0.63) | 0.15 (0.15, 0.16) | 0.11 (0.11, 0.12) | 0.21 (0.20, 0.22) | 0.22 (0.21, 0.23) | 0.20 (0.19, 0.21) | 0.25 (0.24, 0.27) | 0.20 (0.19, 0.21) | 0.19 (0.18, 0.20) |
| | PT | 4.16 (4.11, 4.21) | 1.29 (1.18, 1.41) | 6.42 (6.31, 6.52) | 3.89 (3.72, 4.08) | 6.90 (6.75, 7.05) | 4.23 (3.34, 5.35) | 0.26 (0.18, 0.38) | 0.66 (0.63, 0.68) | 0.89 (0.84, 0.94) | 0.39 (0.36, 0.42) | 0.32 (0.29, 0.35) | 1.71 (1.60, 1.82) | 0.78 (0.73, 0.83) | 0.50 (0.45, 0.55) | 1.71 (1.54, 1.89) | 1.68 (1.50, 1.88) | 0.97 (0.82, 1.15) |
| | LAc | 4.15 (4.10, 4.20) | 1.37 (1.25, 1.51) | 1.26 (1.17, 1.36) | 4.71 (4.50, 4.93) | 3.84 (3.69, 4.01) | 194.06 (177.49, 212.17) | 0.34 (0.24, 0.48) | 0.16 (0.15, 0.18) | 0.20 (0.17, 0.23) | 0.12 (0.10, 0.14) | 0.08 (0.06, 0.10) | 0.28 (0.23, 0.33) | 0.22 (0.19, 0.26) | 0.19 (0.16, 0.23) | 0.35 (0.27, 0.46) | 0.18 (0.12, 0.27) | 0.13 (0.08, 0.22) |
| Specialist | OS | 1.24 (1.21, 1.27) | 0.61 (0.58, 0.65) | 1.72 (1.68, 1.77) | 1.33 (1.28, 1.38) | 1.73 (1.68, 1.78) | 0.87 (0.70, 1.08) | 0.22 (0.19, 0.26) | 1.27 (1.27, 1.28) | 2.46 (2.43, 2.49) | 0.82 (0.80, 0.84) | 0.45 (0.43, 0.46) | 2.52 (2.45, 2.58) | 1.12 (1.10, 1.15) | 0.87 (0.84, 0.89) | 2.20 (2.11, 2.30) | 2.70 (2.59, 2.83) | 1.19 (1.11, 1.27) |
| | PMR | 1.42 (1.38, 1.46) | 0.69 (0.64, 0.75) | 1.91 (1.85, 1.98) | 1.47 (1.39, 1.55) | 1.91 (1.84, 1.99) | 3.76 (3.15, 4.48) | 0.85 (0.74, 0.97) | 0.97 (0.96, 0.99) | 1.32 (1.28, 1.35) | 0.68 (0.65, 0.70) | 0.49 (0.47, 0.51) | 2.27 (2.18, 2.35) | 1.56 (1.52, 1.61) | 1.12 (1.07, 1.17) | 3.97 (3.79, 4.17) | 2.88 (2.71, 3.06) | 0.93 (0.83, 1.04) |
| | PM | 0.44 (0.41, 0.49) | 0.21 (0.17, 0.26) | 0.58 (0.53, 0.64) | 0.38 (0.32, 0.45) | 0.58 (0.52, 0.65) | 0.49 (0.25, 0.94) | 0.22 (0.15, 0.33) | 0.67 (0.65, 0.69) | 0.47 (0.43, 0.51) | 0.43 (0.40, 0.46) | 0.48 (0.44, 0.51) | 1.57 (1.47, 1.68) | 2.35 (2.29, 2.43) | 1.68 (1.60, 1.76) | 6.17 (5.85, 6.50) | 5.19 (4.87, 5.53) | 0.68 (0.55, 0.83) |
| | Neuro | 1.11 (1.06, 1.16) | 0.45 (0.39, 0.51) | 1.57 (1.49, 1.66) | 1.14 (1.05, 1.24) | 1.54 (1.45, 1.64) | 0.43 (0.23, 0.81) | 0.09 (0.05, 0.15) | 1.21 (1.19, 1.22) | 1.92 (1.86, 1.97) | 0.59 (0.56, 0.63) | 0.64 (0.61, 0.68) | 3.94 (3.81, 4.07) | 1.82 (1.76, 1.88) | 1.45 (1.38, 1.53) | 2.50 (2.31, 2.70) | 5.79 (5.48, 6.12) | 3.34 (3.08, 3.63) |
| | Neurologist | 0.85 (0.82, 0.89) | 0.64 (0.58, 0.69) | 1.07 (1.02, 1.13) | 0.90 (0.83, 0.97) | 1.05 (0.99, 1.11) | 0.87 (0.62, 1.22) | 0.15 (0.11, 0.21) | 0.96 (0.94, 0.97) | 0.56 (0.53, 0.59) | 0.62 (0.60, 0.65) | 0.82 (0.79, 0.85) | 2.57 (2.48, 2.66) | 1.21 (1.17, 1.26) | 0.78 (0.74, 0.82) | 2.10 (1.96, 2.24) | 2.36 (2.21, 2.53) | 1.31 (1.18, 1.45) |
| | Rheu | 0.80 (0.75, 0.86) | 0.93 (0.83, 1.03) | 0.89 (0.82, 0.97) | 0.86 (0.76, 0.96) | 0.88 (0.80, 0.97) | 1.07 (0.66, 1.74) | 0.13 (0.08, 0.23) | 1.06 (1.04, 1.09) | 1.28 (1.22, 1.34) | 0.88 (0.84, 0.93) | 0.76 (0.71, 0.80) | 1.26 (1.16, 1.37) | 1.10 (1.04, 1.17) | 0.96 (0.89, 1.03) | 1.88 (1.69, 2.08) | 1.21 (1.05, 1.40) | 0.89 (0.73, 1.07) |
| | MD (Oth) | 1.06 (1.01, 1.10) | 1.55 (1.46, 1.65) | 0.89 (0.84, 0.94) | 1.03 (0.95, 1.11) | 0.85 (0.79, 0.91) | 1.59 (1.21, 2.10) | 0.88 (0.76, 1.02) | 0.69 (0.67, 0.70) | 0.71 (0.68, 0.74) | 0.69 (0.66, 0.72) | 0.50 (0.47, 0.52) | 0.94 (0.88, 1.01) | 1.32 (1.28, 1.37) | 1.33 (1.28, 1.39) | 0.97 (0.88, 1.08) | 1.08 (0.97, 1.20) | 1.55 (1.40, 1.72) |
| Emergency/ Urgent Care | EM | 0.55 (0.53, 0.58) | 0.60 (0.56, 0.65) | 0.62 (0.59, 0.66) | 0.65 (0.60, 0.70) | 0.63 (0.59, 0.67) | 0.53 (0.37, 0.76) | 0.10 (0.07, 0.14) | 1.08 (1.06, 1.09) | 1.33 (1.30, 1.37) | 1.07 (1.05, 1.10) | 1.24 (1.21, 1.26) | 0.68 (0.64, 0.73) | 1.84 (1.80, 1.89) | 0.99 (0.95, 1.03) | 0.57 (0.52, 0.64) | 0.66 (0.59, 0.73) | 8.54 (8.18, 8.90) |
| | Rad | 0.20 (0.18, 0.21) | 0.09 (0.07, 0.11) | 0.27 (0.25, 0.30) | 0.20 (0.17, 0.23) | 0.27 (0.24, 0.30) | 0.24 (0.14, 0.42) | 0.02 (0.01, 0.04) | 1.21 (1.20, 1.23) | 2.19 (2.15, 2.23) | 0.06 (0.05, 0.07) | 0.05 (0.05, 0.06) | 2.40 (2.32, 2.48) | 1.13 (1.09, 1.16) | 0.08 (0.07, 0.10) | 0.49 (0.44, 0.55) | 0.57 (0.50, 0.64) | 6.74 (6.44, 7.06) |
| | UC | 0.65 (0.58, 0.72) | 0.74 (0.63, 0.87) | 0.62 (0.54, 0.71) | 0.66 (0.55, 0.79) | 0.72 (0.62, 0.83) | 0.56 (0.23, 1.34) | 0.44 (0.29, 0.65) | 1.00 (0.97, 1.03) | 1.16 (1.08, 1.24) | 0.87 (0.81, 0.94) | 1.16 (1.10, 1.23) | 0.64 (0.55, 0.75) | 0.65 (0.59, 0.73) | 0.61 (0.54, 0.70) | 0.54 (0.42, 0.71) | 0.55 (0.41, 0.73) | 0.94 (0.73, 1.20) |

PCP=Primary Care Provider, PA=Physician Assistant, DO=Doctor of Osteopathy, DC=Doctor of Chiropractic, PT=Physical Therapist, LAc=Licensed Acupuncturist, OS=Orthpedic Surgeon, PMR=Physical Medicine & Rehabilitation, PM=Pain Management, NS=Neurosurgeon, Neuro=Neurologist, Rheum=Rheumatologist, MD Oth=Other MD specialty, EM=Emergency Medicine, Rad=Radiologist, UC=Urgent Care

Cells in red are not different than the PCP reference (p=.05)
