## Supplement 5 - Non-surgical Timing for "Neck pain care pathways and costs: association with the type of initial contact health care provider. A retrospective cohort study"

**Supplement 5 - Neck pain # of days into episode when service initially provided by type of initial contact health care provider (HCP)**

| All Non-Surgical Episodes Median (Q1, Q3) | | First Line: Any | Manipulation - Chiropractic | Active Care | Passive Therapy | Manual Therapy | Acupuncture | Manipulation - Osteopathic |
|---|---|---|---|---|---|---|---|---|
| Primary Care | PCP | 19 (3, 63) | 41 (11, 101) | 22 (7, 64) | 33 (9, 88) | 23 (7, 64) | 31 (1, 98) | 0 (0, 0) |
| | Nurse | 19 (3, 61) | 26 (5, 78) | 20 (5, 58) | 23 (5, 71) | 21 (6, 62) | 44 (14, 95) | 5 (0, 28) |
| | PA | 21 (7, 63) | 33 (8, 85) | 20 (7, 57) | 29 (9, 78) | 21 (8, 60) | 54 (28, 117) | 19 (6, 80) |
| | DO | 0 (0, 0) | 69 (9, 119) | 8 (0, 56) | 0 (0, 29) | 36 (9, 84) | 36 (18, 76) | 0 (0, 0) |
| | All | 19 (3, 62) | 38 (9, 95) | 22 (7, 63) | 30 (8, 84) | 23 (7, 63) | 34 (3, 100) | 0 (0, 0) |
| Non-Prescriber | DC | 0 (0, 0) | 0 (0, 0) | 0 (0, 4) | 0 (0, 1) | 0 (0, 7) | 7 (0, 61) | 45 (5, 128) |
| | PT | 0 (0, 0) | 41 (10, 60) | 0 (0, 1) | 4 (0, 22) | 0 (0, 6) | 18 (0, 94) | 33 (8, 106) |
| | LAc | 0 (0, 0) | 35 (4, 58) | 3 (0, 38) | 0 (0, 0) | 0 (0, 1) | 0 (0, 0) | 19 (1, 49) |
| | All | 0 (0, 0) | 0 (0, 0) | 0 (0, 4) | 0 (0, 1) | 0 (0, 7) | 0 (0, 0) | 40 (5, 105) |
| Specialist | OS | 14 (6, 40) | 42 (13, 105) | 14 (6, 37) | 20 (8, 50) | 15 (7, 39) | 56 (16, 118) | 0 (0, 43) |
| | PMR | 10 (0, 32) | 26 (3, 96) | 12 (2, 33) | 17 (2, 46) | 14 (3, 36) | 0 (0, 36) | 0 (0, 23) |
| | PM | 2 (0, 34) | 4 (0, 68) | 6 (0, 34) | 19 (0, 49) | 12 (0, 44) | 77 (29, 107) | 0 (0, 0) |
| | NS | 29 (12, 63) | 64 (22, 153) | 28 (12, 58) | 35 (15, 71) | 30 (13, 58) | 72 (30, 273) | 99 (79, 185) |
| | Neuro | 32 (12, 84) | 58 (24, 134) | 31 (12, 81) | 43 (18, 100) | 31 (12, 80) | 85 (26, 121) | 54 (0, 138) |
| | Rheu | 45 (15, 107) | 62 (20, 126) | 47 (15, 120) | 69 (22, 132) | 46 (17, 112) | 20 (12, 50) | 12 (3, 54) |
| | MD (Oth) | 41 (10, 98) | 49 (17, 109) | 55 (20, 122) | 56 (21, 118) | 55 (19, 130) | 59 (28, 91) | 0 (0, 0) |
| | All | 18 (5, 54) | 46 (14, 111) | 18 (6, 50) | 27 (9, 70) | 20 (7, 52) | 32 (0, 91) | 0 (0, 31) |
| Emergency / Urgent Care | EM | 20 (7, 47) | 18 (6, 57) | 23 (11, 50) | 23 (9, 51) | 25 (11, 48) | 22 (6, 56) | 11 (1, 68) |
| | Rad | 18 (6, 40) | 29 (3, 105) | 20 (8, 39) | 22 (6, 46) | 21 (8, 42) | 32 (18, 81) | 90 (31, 102) |
| | UC | 18 (5, 38) | 20 (6, 48) | 22 (10, 38) | 22 (7, 42) | 21 (10, 36) | 29 (13, 60) | 0 (0, 14) |
| | All | 19 (7, 45) | 19 (6, 57) | 22 (10, 45) | 23 (8, 49) | 23 (10, 45) | 24 (9, 60) | 7 (0, 51) |

| All Non-Surgical Episodes Median (Q1, Q3) | | Second Line | | | | | Third Line | | | | |
|---|---|---|---|---|---|---|---|---|---|---|---|
| | | Any | Imaging - Radiography | Rx - NSAID | Rx - Skeletal Muscle Relaxant | Imaging - MRI | Any | Rx-Opioid | Spinal Injection | Spinal Surgery | Imaging-CT |
| Primary Care | PCP | 0 (0, 0) | 0 (0, 31) | 0 (0, 11) | 0 (0, 1) | 30 (9, 93) | 0 (0, 41) | 0 (0, 37) | 27 (0, 107) | N/A | 9 (0, 63) |
| | Nurse | 0 (0, 0) | 0 (0, 21) | 0 (0, 7) | 0 (0, 0) | 31 (8, 86) | 1 (0, 36) | 2 (0, 48) | 10 (0, 58) | N/A | 0 (0, 30) |
| | PA | 0 (0, 0) | 0 (0, 7) | 0 (0, 3) | 0 (0, 0) | 21 (6, 70) | 0 (0, 26) | 0 (0, 32) | 16 (0, 75) | N/A | 0 (0, 13) |
| | DO | 5 (0, 47) | 7 (0, 52) | 30 (0, 101) | 4 (0, 38) | 24 (10, 65) | 7 (0, 46) | 24 (2, 74) | 0 (0, 36) | N/A | 0 (0, 26) |
| | All | 0 (0, 0) | 0 (0, 27) | 0 (0, 9) | 0 (0, 0) | 29 (8, 89) | 0 (0, 38) | 0 (0, 39) | 20 (0, 91) | N/A | 3 (0, 51) |
| Non-Prescriber | DC | 0 (0, 39) | 0 (0, 1) | 78 (26, 156) | 55 (13, 135) | 55 (19, 128) | 57 (15, 138) | 67 (22, 153) | 38 (7, 107) | N/A | 38 (1, 121) |
| | PT | 17 (0, 56) | 13 (0, 56) | 49 (14, 123) | 38 (8, 90) | 35 (5, 73) | 26 (3, 78) | 32 (8, 94) | 38 (8, 99) | N/A | 8 (0, 64) |
| | LAc | 42 (10, 114) | 44 (14, 102) | 62 (18, 155) | 46 (9, 154) | 60 (18, 160) | 61 (18, 151) | 82 (25, 173) | 58 (14, 96) | N/A | 28 (14, 106) |
| | All | 0 (0, 42) | 0 (0, 2) | 76 (25, 155) | 52 (13, 134) | 50 (16, 120) | 54 (14, 135) | 65 (21, 150) | 38 (7, 105) | N/A | 33 (0, 115) |
| Specialist | OS | 0 (0, 0) | 0 (0, 0) | 0 (0, 28) | 0 (0, 35) | 10 (2, 30) | 7 (0, 57) | 8 (0, 65) | 20 (0, 67) | N/A | 15 (0, 81) |
| | PMR | 0 (0, 7) | 0 (0, 0) | 4 (0, 64) | 0 (0, 46) | 10 (1, 38) | 2 (0, 39) | 2 (0, 42) | 14 (0, 67) | N/A | 24 (0, 96) |
| | PM | 0 (0, 21) | 0 (0, 34) | 3 (0, 63) | 0 (0, 28) | 13 (1, 48) | 0 (0, 13) | 0 (0, 11) | 0 (0, 36) | N/A | 45 (1, 109) |
| | NS | 0 (0, 7) | 0 (0, 14) | 31 (2, 107) | 14 (0, 79) | 3 (0, 18) | 15 (0, 64) | 19 (1, 80) | 50 (16, 94) | N/A | 14 (0, 60) |
| | Neuro | 0 (0, 16) | 23 (0, 93) | 14 (0, 93) | 0 (0, 20) | 9 (0, 29) | 4 (0, 64) | 19 (0, 97) | 0 (0, 55) | N/A | 8 (0, 63) |
| | Rheu | 0 (0, 6) | 0 (0, 44) | 0 (0, 39) | 0 (0, 36) | 47 (10, 146) | 14 (0, 78) | 15 (0, 83) | 13 (0, 90) | N/A | 48 (14, 133) |
| | MD (Oth) | 2 (0, 68) | 32 (0, 106) | 2 (0, 85) | 11 (0, 86) | 50 (9, 127) | 0 (0, 28) | 0 (0, 23) | 55 (3, 137) | N/A | 0 (0, 60) |
| | All | 0 (0, 3) | 0 (0, 0) | 1 (0, 53) | 0 (0, 41) | 10 (0, 36) | 2 (0, 46) | 3 (0, 53) | 15 (0, 70) | N/A | 14 (0, 76) |
| Emergency / Urgent Care | EM | 0 (0, 0) | 0 (0, 0) | 0 (0, 2) | 0 (0, 1) | 15 (0, 49) | 0 (0, 0) | 0 (0, 4) | 4 (0, 62) | N/A | 0 (0, 0) |
| | Rad | 0 (0, 0) | 0 (0, 0) | 10 (0, 63) | 1 (0, 25) | 0 (0, 0) | 0 (0, 0) | 19 (1, 71) | 34 (10, 70) | N/A | 0 (0, 0) |
| | UC | 0 (0, 0) | 0 (0, 0) | 0 (0, 0) | 0 (0, 0) | 29 (9, 76) | 0 (0, 16) | 0 (0, 21) | 14 (0, 76) | N/A | 0 (0, 1) |
| | All | 0 (0, 0) | 0 (0, 0) | 0 (0, 2) | 0 (0, 1) | 0 (0, 9) | 0 (0, 0) | 0 (0, 7) | 20 (0, 68) | N/A | 0 (0, 0) |

PCP=Primary Care Provider, PA=Physician Assistant, DO=Doctor of Osteopathy, DC=Doctor of Chiropractic, PT=Physical Therapist, LAc=Licensed Acupuncturist, OS=Orthpedic Surgeon, PMR=Physical Medicine & Rehabilitation, PM=Pain Management, NS=Neurosurgeon, Neuro=Neurologist, Rheum=Rheumatologist, MD Oth=Other MD specialty, EM=Emergency Medicine, Rad=Radiologist, UC=Urgent Care
