## Supplement 6 - Pooled Timing for "Neck pain care pathways and costs: association with the type of initial contact health care provider. A retrospective cohort study"

### Supplement 6 - Neck pain # of days into episode when service initially provided by type of initial contact health care provider (HCP)

| Combined Surgical and Non-Surgical (Pooled) Episodes<br>Median (Q1, Q3) | | First Line | | | | | | |
|---|---|---|---|---|---|---|---|---|
| | | Any | Manipulation - Chiropractic | Active Care | Passive Therapy | Manual Therapy | Acupuncture | Manipulation - Osteopathic |
| Primary Care | PCP | 21 (3, 68) | 42 (11, 104) | 26 (8, 73) | 35 (10, 92) | 26 (8, 73) | 34 (2, 105) | 0 (0, 0) |
| | Nurse | 21 (4, 67) | 27 (6, 81) | 22 (6, 68) | 25 (6, 77) | 24 (7, 72) | 44 (14, 95) | 6 (0, 34) |
| | PA | 22 (7, 67) | 33 (8, 86) | 22 (7, 65) | 30 (10, 81) | 25 (9, 66) | 58 (27, 132) | 25 (7, 78) |
| | DO | 0 (0, 0) | 76 (27, 124) | 11 (0, 66) | 0 (0, 32) | 36 (12, 90) | 18 (0, 56) | 0 (0, 0) |
| | All | 21 (4, 67) | 38 (9, 98) | 25 (7, 71) | 33 (9, 88) | 26 (8, 72) | 36 (4, 105) | 0 (0, 0) |
| Non-Prescriber | DC | 0 (0, 0) | 0 (0, 0) | 0 (0, 5) | 0 (0, 1) | 0 (0, 7) | 8 (0, 62) | 45 (6, 134) |
| | PT | 0 (0, 0) | 44 (11, 68) | 0 (0, 2) | 5 (0, 24) | 0 (0, 7) | 20 (0, 94) | 34 (8, 104) |
| | LAc | 0 (0, 0) | 34 (4, 58) | 3 (0, 38) | 0 (0, 0) | 0 (0, 1) | 0 (0, 0) | 19 (1, 49) |
| | All | 0 (0, 0) | 0 (0, 0) | 0 (0, 5) | 0 (0, 1) | 0 (0, 7) | 0 (0, 0) | 41 (6, 109) |
| Specialist | OS | 17 (6, 49) | 43 (14, 107) | 17 (7, 48) | 22 (8, 58) | 18 (7, 49) | 63 (28, 118) | 0 (0, 53) |
| | PMR | 13 (1, 38) | 34 (4, 102) | 14 (3, 39) | 20 (3, 55) | 15 (4, 41) | 0 (0, 41) | 0 (0, 28) |
| | PM | 7 (0, 49) | 30 (0, 92) | 10 (0, 50) | 20 (2, 62) | 17 (0, 53) | 77 (38, 137) | 0 (0, 5) |
| | NS | 38 (14, 88) | 67 (22, 156) | 39 (14, 86) | 46 (16, 98) | 42 (16, 94) | 72 (30, 273) | 132 (82, 237) |
| | Neuro | 37 (13, 94) | 60 (26, 134) | 37 (13, 95) | 46 (19, 101) | 35 (14, 94) | 85 (30, 136) | 52 (2, 130) |
| | Rheu | 51 (16, 119) | 62 (20, 126) | 55 (18, 133) | 71 (22, 142) | 55 (18, 122) | 21 (14, 55) | 12 (3, 54) |
| | MD (Oth) | 43 (11, 105) | 51 (18, 112) | 59 (22, 129) | 60 (22, 126) | 58 (21, 133) | 59 (28, 99) | 0 (0, 0) |
| | All | 21 (6, 64) | 49 (14, 114) | 22 (7, 63) | 29 (10, 78) | 22 (7, 64) | 37 (2, 95) | 0 (0, 35) |
| Emergency/ Urgent Care | EM | 20 (8, 49) | 17 (6, 57) | 24 (11, 51) | 24 (10, 53) | 26 (11, 50) | 23 (6, 56) | 9 (1, 63) |
| | Rad | 20 (7, 41) | 30 (4, 112) | 20 (8, 41) | 23 (7, 54) | 22 (9, 43) | 33 (19, 91) | 60 (26, 99) |
| | UC | 18 (6, 42) | 20 (6, 50) | 24 (11, 44) | 24 (7, 43) | 23 (10, 38) | 29 (13, 60) | 0 (0, 14) |
| | All | 20 (7, 46) | 19 (6, 58) | 23 (10, 48) | 24 (8, 53) | 24 (11, 48) | 27 (10, 60) | 7 (0, 46) |

| Combined Surgical and Non-Surgical (Pooled) Episodes<br>Median (Q1, Q3) | | Second Line | | | | | Third Line | | | | |
|---|---|---|---|---|---|---|---|---|---|---|---|
| | | Any | Imaging - Radiography | Rx - NSAID | Rx - Skeletal Muscle Relaxant | Imaging - MRI | Any | Rx-Opioid | Spinal Injection | Spinal Surgery | Imaging-CT |
| Primary Care | PCP | 0 (0, 1) | 1 (0, 40) | 0 (0, 15) | 0 (0, 2) | 31 (9, 95) | 2 (0, 54) | 1 (0, 49) | 48 (10, 126) | 64 (28, 132) | 15 (0, 78) |
| | Nurse | 0 (0, 0) | 0 (0, 29) | 0 (0, 11) | 0 (0, 0) | 30 (8, 88) | 4 (0, 45) | 4 (0, 58) | 28 (2, 84) | 46 (16, 106) | 2 (0, 44) |
| | PA | 0 (0, 0) | 0 (0, 13) | 0 (0, 7) | 0 (0, 0) | 21 (6, 67) | 1 (0, 39) | 1 (0, 42) | 34 (6, 94) | 50 (19, 105) | 0 (0, 25) |
| | DO | 4 (0, 48) | 9 (0, 76) | 20 (0, 99) | 4 (0, 45) | 26 (10, 69) | 10 (0, 54) | 29 (0, 80) | 18 (0, 59) | 78 (39, 110) | 26 (0, 99) |
| | All | 0 (0, 0) | 0 (0, 35) | 0 (0, 14) | 0 (0, 1) | 29 (8, 91) | 2 (0, 51) | 1 (0, 50) | 42 (7, 113) | 59 (24, 126) | 8 (0, 67) |
| Non-Prescriber | DC | 0 (0, 41) | 0 (0, 2) | 77 (26, 156) | 55 (13, 136) | 52 (18, 127) | 57 (16, 138) | 67 (21, 153) | 53 (15, 128) | 78 (34, 158) | 41 (2, 131) |
| | PT | 19 (0, 56) | 18 (0, 58) | 50 (16, 122) | 42 (10, 108) | 34 (7, 74) | 34 (7, 87) | 41 (11, 100) | 45 (16, 109) | 57 (21, 114) | 22 (0, 86) |
| | LAc | 41 (10, 112) | 43 (13, 101) | 61 (19, 145) | 47 (12, 147) | 59 (19, 170) | 61 (18, 142) | 82 (24, 169) | 58 (14, 104) | 42 (23, 135) | 37 (23, 199) |
| | All | 0 (0, 43) | 0 (0, 4) | 74 (24, 154) | 52 (13, 135) | 49 (16, 120) | 54 (15, 133) | 65 (21, 150) | 52 (15, 125) | 74 (31, 149) | 39 (1, 128) |
| Specialist | OS | 0 (0, 0) | 0 (0, 0) | 0 (0, 36) | 2 (0, 56) | 10 (2, 32) | 19 (0, 63) | 20 (0, 79) | 35 (10, 81) | 44 (18, 87) | 28 (1, 99) |
| | PMR | 0 (0, 11) | 0 (0, 10) | 8 (0, 74) | 2 (0, 63) | 11 (2, 41) | 7 (0, 42) | 7 (0, 62) | 21 (0, 64) | 26 (6, 68) | 39 (2, 110) |
| | PM | 0 (0, 27) | 0 (0, 59) | 7 (0, 69) | 0 (0, 33) | 12 (0, 52) | 0 (0, 18) | 0 (0, 18) | 6 (0, 35) | 6 (0, 33) | 52 (8, 145) |
| | NS | 0 (0, 11) | 2 (0, 41) | 42 (5, 121) | 31 (1, 94) | 4 (0, 21) | 22 (1, 62) | 33 (5, 82) | 46 (19, 98) | 40 (14, 83) | 24 (0, 82) |
| | Neuro | 0 (0, 18) | 30 (0, 102) | 18 (0, 98) | 0 (0, 27) | 10 (0, 33) | 6 (0, 61) | 29 (0, 105) | 21 (0, 91) | 22 (0, 78) | 21 (0, 91) |
| | Rheu | 0 (0, 9) | 0 (0, 54) | 0 (0, 42) | 0 (0, 42) | 49 (12, 147) | 18 (0, 85) | 19 (0, 96) | 43 (0, 120) | 83 (31, 172) | 67 (15, 143) |
| | MD (Oth) | 3 (0, 68) | 35 (0, 113) | 5 (0, 88) | 14 (0, 91) | 55 (11, 134) | 0 (0, 41) | 0 (0, 32) | 71 (18, 156) | 68 (23, 147) | 8 (0, 75) |
| | All | 0 (0, 6) | 0 (0, 11) | 3 (0, 62) | 2 (0, 57) | 10 (0, 39) | 9 (0, 53) | 11 (0, 70) | 27 (0, 78) | 34 (8, 81) | 27 (0, 98) |
| Emergency/ Urgent Care | EM | 0 (0, 0) | 0 (0, 0) | 0 (0, 2) | 0 (0, 1) | 15 (1, 49) | 0 (0, 0) | 0 (0, 6) | 34 (2, 87) | 34 (7, 82) | 0 (0, 0) |
| | Rad | 0 (0, 0) | 0 (0, 0) | 14 (0, 77) | 4 (0, 46) | 0 (0, 0) | 0 (0, 0) | 28 (2, 82) | 29 (10, 66) | 27 (10, 60) | 0 (0, 0) |
| | UC | 0 (0, 0) | 0 (0, 0) | 0 (0, 1) | 0 (0, 0) | 30 (10, 77) | 0 (0, 29) | 1 (0, 29) | 35 (1, 86) | 59 (25, 128) | 0 (0, 12) |
| | All | 0 (0, 0) | 0 (0, 0) | 0 (0, 3) | 0 (0, 1) | 0 (0, 12) | 0 (0, 0) | 1 (0, 10) | 32 (4, 77) | 31 (8, 77) | 0 (0, 0) |

PCP=Primary Care Provider, PA=Physician Assistant, DO=Doctor of Osteopathy, DC=Doctor of Chiropractic, PT=Physical Therapist, LAc=Licensed Acupuncturist, OS=Orthpedic Surgeon, PMR=Physical Medicine & Rehabilitation, PM=Pain Management, NS=Neurosurgeon, Neuro=Neurologist, Rheum=Rheumatologist, MD Oth=Other MD specialty, EM=Emergency Medicine, Rad=Radiologist, UC=Urgent Care
