## Supplement 7 - Pooled Cost for "Neck pain care pathways and costs: association with the type of initial contact health care provider. A retrospective cohort study"

| Supplement 7 - Neck pain total episode cost by episode sequence cohort | | | | | | |
|---|---|---|---|---|---|---|
| Combined Surgical and Non-surgical (Pooled) Episodes Median (IQR) (Q1, Q3) | | All | Single episode | Multiple episodes | | |
| | | | | First | Second | Third+ |
| Primary Care | PCP | $165 (57, 563) | $168 (63, 564) | $209 (68, 813) | $125 (26, 404) | $97 (21, 266) |
| | Nurse | $167 (40, 688) | $174 (48, 704) | $231 (55, 1036) | $106 (18, 426) | $95 (21, 275) |
| | PA | $222 (57, 884) | $229 (66, 887) | $300 (79, 1349) | $141 (20, 607) | $112 (19, 403) |
| | DO | $234 (107, 574) | $260 (107, 641) | $233 (84, 539) | $219 (116, 487) | $71 (69, 113) |
| | All | $170 (55, 606) | $174 (62, 610) | $218 (67, 883) | $124 (24, 424) | $97 (21, 274) |
| Non-Prescriber | DC | $165 (65, 405) | $180 (72, 457) | $165 (68, 399) | $130 (59, 312) | $90 (50, 180) |
| | PT | $700 (343, 1569 | $684 (330, 1603 | $768 (417, 1684 | $700 (370, 1478 | $660 (307, 1119 |
| | LAc | $331 (154, 719) | $331 (154, 719) | $395 (158, 910) | $321 (148, 630) | $276 (160, 552) |
| | All | $172 (66, 433) | $195 (79, 495) | $176 (70, 420) | $135 (60, 330) | $90 (50, 189) |
| Specialist | OS | $397 (147, 1324 | $381 (146, 1269 | $641 (194, 2121 | $310 (117, 1041 | $255 (106, 847) |
| | PMR | $623 (203, 1750 | $619 (214, 1747 | $799 (229, 2248 | $519 (151, 1442 | $327 (91, 1152) |
| | PM | $419 (131, 1318 | $451 (140, 1409 | $430 (138, 1337 | $330 (107, 1113 | $230 (87, 686) |
| | NS | $1110 (313, 3937 | $1208 (348, 4085 | $1570 (433, 6637 | $624 (210, 2271 | $422 (134, 1378 |
| | Neuro | $435 (134, 1319 | $515 (156, 1432 | $504 (148, 1501 | $274 (90, 878) | $174 (71, 319) |
| | Rheu | $278 (73, 969) | $272 (67, 1005) | $349 (108, 1235 | $257 (75, 812) | $162 (39, 477) |
| | MD (Oth) | $154 (28, 664) | $166 (31, 752) | $198 (37, 930) | $104 (17, 369) | $91 (22, 279) |
| | All | $418 (133, 1422 | $432 (139, 1438 | $569 (165, 1948 | $296 (99, 1037) | $200 (72, 563) |
| Emergency/ Urgent Care | EM | $900 (242, 2051 | $927 (255, 2056 | $928 (260, 2286 | $433 (58, 1724) | $338 (18, 1298) |
| | Rad | $191 (69, 683) | $182 (68, 633) | $276 (83, 1044) | $227 (70, 769) | $234 (60, 900) |
| | UC | $209 (110, 405) | $205 (113, 382) | $256 (104, 674) | $248 (86, 557) | $120 (18, 364) |
| | All | $381 (110, 1392 | $385 (111, 1400 | $494 (117, 1586 | $294 (69, 1041) | $246 (40, 959) |

PCP=Primary Care Provider, PA=Physician Assistant, DO=Doctor of Osteopathy, DC=Doctor of Chiropractic, PT=Physical Therapist, LAc=Licensed Acupuncturist, OS=Orthpedic Surgeon, PMR=Physical Medicine & Rehabilitation, PM=Pain Management, NS=Neurosurgeon, Neuro=Neurologist, Rheum=Rheumatologist, MD Oth=Other MD specialty, EM=Emergency Medicine, Rad=Radiologist, UC=Urgent Care
