## Supplement 8 - Pooled Mixed Effects Model for "Neck pain care pathways and costs: association with the type of initial contact health care provider. A retrospective cohort study"

| Supplement 8 - Neck pain mixed effects model - Pooled Sample | | | |
|---|---|---|---|
| Features | Total Episode Cost | % of Episodes With Opioid | % of Episodes With NSAID |
| PCP - (Intercept) | 772 (723, 820) | 0.148 (0.145, 0.150) | 0.306 (0.303, 0.309) |
| Nurse | 41 (-53, 134) | -0.012 (-0.017, -0.007) | 0.010 (0.004, 0.017) |
| PA | 359 (249, 469) | -0.002 (-0.008, 0.004) | 0.002 (-0.005, 0.009) |
| DO | -368 (-990, 254) | -0.108 (-0.146, -0.070) | -0.240 (-0.285, -0.194) |
| DC | -473 (-530, -415) | -0.129 (-0.133, -0.126) | -0.261 (-0.265, -0.257) |
| PT | 786 (583, 989) | -0.100 (-0.111, -0.089) | -0.200 (-0.214, -0.187) |
| LAc | -154 (-388, 80) | -0.132 (-0.145, -0.119) | -0.277 (-0.293, -0.261) |
| OS | 1565 (1468, 1663) | -0.036 (-0.041, -0.030) | -0.053 (-0.060, -0.046) |
| PMR | 1143 (994, 1292) | 0.015 (0.006, 0.024) | -0.112 (-0.123, -0.101) |
| PM | 22 (-190, 233) | 0.087 (0.075, 0.100) | -0.196 (-0.210, -0.181) |
| NS | 5924 (5733, 6115) | 0.044 (0.033, 0.055) | -0.144 (-0.157, -0.130) |
| Neuro | 946 (799, 1094) | -0.061 (-0.069, -0.052) | -0.131 (-0.141, -0.120) |
| Rheum | -69 (-292, 154) | -0.041 (-0.054, -0.028) | -0.042 (-0.057, -0.027) |
| MD (Oth) | 172 (27, 317) | 0.044 (0.036, 0.051) | -0.105 (-0.115, -0.096) |
| EM | 803 (682, 924) | 0.003 (-0.004, 0.009) | 0.018 (0.010, 0.026) |
| Rad | -584 (-710, -457) | -0.183 (-0.190, -0.176) | -0.309 (-0.317, -0.300) |
| UC | -62 (-371, 246) | -0.046 (-0.064, -0.028) | -0.015 (-0.036, 0.006) |
| Individual Age | 14 (12, 16) | 0.001 (0.001, 0.001) | 0.001 (0.000, 0.001) |
| Individual Gender | -300 (-339, -261) | -0.006 (-0.008, -0.004) | 0.003 (0.000, 0.005) |
| ERG® Risk Score | 227 (220, 234) | 0.014 (0.013, 0.014) | 0.004 (0.003, 0.004) |

PCP=Primary Care Provider, PA=Physician Assistant, DO=Doctor of Osteopathy, DC=Doctor of Chiropractic, PT=Physical Therapist, LAc=Licensed Acupuncturist, OS=Orthpedic Surgeon, PMR=Physical Medicine & Rehabilitation, PM=Pain Management, NS=Neurosurgeon, Neuro=Neurologist, Rheum=Rheumatologist, MD Oth=Other MD specialty, EM=Emergency Medicine, Rad=Radiologist, UC=Urgent Care

Pooled = Combined surgical and non-surgical episodes

Values in red were not statistically significant (p > 0.05) compared to intercept
Values in black were statistically significant (p > 0.05) compared to intercept
